# COVID-19 Open Source Data Sets: A Comprehensive Survey

**DOI:** 10.1101/2020.05.19.20107532

**Authors:** Junaid Shuja, Eisa Alanazi, Waleed Alasmary, Abdulaziz Alashaikh

## Abstract

In December 2019, a novel virus named COVID-19 emerged in the city of Wuhan, China. In early 2020, the COVID-19 virus spread in all continents of the world except Antarctica causing widespread infections and deaths due to its contagious characteristics and no medically proven treatment. The COVID-19 pandemic has been termed as the most consequential global crisis after the World Wars. The first line of defense against the COVID-19 spread are the non-pharmaceutical measures like social distancing and personal hygiene. The great pandemic affecting billions of lives economically and socially has motivated the scientific community to come up with solutions based on computer-aided digital technologies for diagnosis, prevention, and estimation of COVID-19. Some of these efforts focus on statistical and Artificial Intelligence-based analysis of the available data concerning COVID-19. All of these scientific efforts necessitate that the data brought to service for the analysis should be open source to promote the extension, validation, and collaboration of the work in the fight against the global pandemic. Our survey is motivated by the open source efforts that can be mainly categorized as **(a)** COVID-19 diagnosis from CT scans, X-ray images, and cough sounds, **(b)** COVID-19 case reporting, transmission estimation, and prognosis from epidemiological, demographic, and mobility data, **(c)** COVID-19 emotional and sentiment analysis from social media, and **(d)** knowledge-based discovery and semantic analysis from the collection of scholarly articles covering COVID-19. We survey and compare research works in these directions that are accompanied by open source data and code. Future research directions for data-driven COVID-19 research are also debated. We hope that the article will provide the scientific community with an initiative to start open source extensible and transparent research in the collective fight against the COVID-19 pandemic.

## Introduction

The COVID-19 virus has been declared a pandemic by the World Health Organization (WHO) with more than ten million cases and 503862 deaths across the world as per WHO statistics of 30 June 2020 [1]. COVID-19 is caused by Severe acute respiratory syndrome Coronavirus 2 (SARS-CoV-2) and was declared pandemic by WHO on March 11, 2020. The cure to COVID-19 can take several months due to its clinical trials on humans of varying ages and ethnicity before approval. The cure to COVID-19 can be further delayed due to possible genetic mutations shown by the virus [2]. The pandemic situation is affecting billions of people socially, economically, and medically with drastic changes in social relationships, health policies, trade, work and educational environments. The global pandemic is a threat to human society and calls for immediate actions. The COVID-19 pandemic has motivated the research community to aid front-line medical service staff with cutting edge research for mitigation, detection, and prevention of the virus [3].

Scientific community has brainstormed to come up with ideas that can limit the crisis and help prevent future such pandemics. Other than medical science researchers and virology specialists, scientists supported with digital technologies have tackled the pandemic with novel methods. Two significant scientific communities aided with digital technologies can be identified in the fight against COVID-19. The main digital effort in this regard comes from the Artificial Intelligence (AI) community in the form of automated COVID-19 detection from Computed Tomography (CT) scans and X-ray images. The second such community aided by digital technologies is of mathematicians and epidemiologists who are developing complex virus diffusion and transmission models to estimate virus spread under various mobility and social distancing scenarios [4]. Besides these two major scientific communities, efforts are being made for analyzing social and emotional behavior from social media [5], collecting scholarly articles for knowledge-based discovery [6], detection COVID-19 from cough samples [7], and automated contact tracing [2].

Artificial Intelligence (AI) and Machine Learning (ML) techniques have been prominently used to efficiently solve various computer science problems ranging from bio-informatics to image processing. ML is based on the premise that an intelligent machine should be able to learn and adapt from its environment based on its experiences without explicit programming [8, 9]. ML models and algorithms have been standardized across multiple programming languages such as, Python and R. The main challenge to the application of ML models is the availability of the open source data [10, 11]. Given publicly available data sets, ML techniques can aid the fight against COVID-19 on multiple fronts. The principal such application is ML based COVID-19 diagnosis from CT scans and X-rays that can lower the burden on short supplies of reverse transcriptase polymerase chain reaction (RT-PCR) test kits [12, 13]. Similarly, statistical and epidemiological analysis of COVID-19 case reports can help find a relation between human mobility and virus transmission. Moreover, social media data mining can provide sentiment and socio-economic analysis in current pandemic for policy makers. Therefore, the COVID-19 pandemic has necessitated collection of new data sets regarding human mobility, epidemiology, psychology, and radiology to aid scientific efforts [14]. It must be noted that while digital technologies are aiding in the combat against COVID-19, they are also being utilized for spread of misinformation [15], hatred [16], propaganda [17], and online financial scams [18].

Data is an essential element for the efficient implementation of scientific methods. Two approaches are followed by the research community while performing scientific research. The research methods and data are either closed-source to protect proprietary scientific contributions or open source. The open source research leads to higher usability, verifiability, transparency, quality, and collaborative research [19, 20]. In the existing COVID-19 pandemic, the open source approach is deemed more effective for mitigation and detection of the COVID-19 virus due to its aforementioned characteristics. Specifically, open source COVID-19 diagnosis techniques are necessary to gain trust of medical staff and patients while engaging the research community across the globe. We emphasize that the COVID-19 pandemic demands a unified and collaborative approach with open source data and methodology so that the scientific community across the globe can join hands with verifiable and transparent research [21, 22].

The combination of AI and open source data sets produces a practical solution for COVID-19 diagnosis that can be implemented in hospitals worldwide. Automated CT scan based COVID-19 detection techniques work with training the learning model on existing CT scan data sets that contain labeled images of COVID-19 positive and normal cases. Similarly, the detection of COVID-19 from cough requires both normal and infected samples to learn and distinguish features of the infected person from a healthy person. Therefore, it is necessary to provide open source data sets and methods so that **(a)** researchers across globe can enhance and modify existing work to limit the global pandemic, **(b)** existing techniques are verified for correctness by researchers across the board before implementation in real-world scenarios, and **(c)** researchers collaborate to aggregate data sets and enhance the performance of AI/ML methods in community-oriented research and development [23, 24, 25]. The fruits of open source science can be seen in abundance among the community. Some of the leading hospitals across the world are utilizing AI/ML algorithms to diagnose COVID-19 cases from CT scans/X-ray images after preliminary trails of the technology ^2^.

Efforts have been made on surveying the role of ICT in combating COVID-19 pandemic [26, 27]. Specifically, the role of AI, data science, and big data in the management of COVID-19 has been surveyed. Researchers [13] surveyed AI-based techniques for data acquisition, segmentation, and diagnosis for COVID-19. The article was not focused on works that are accompanied by publicly available data sets. Moreover, the article focused only on the applications of ML towards medical diagnosis. Authors [14] listed publicly available medical data sets for COVID-19. The work did not detail the AI applications of the data set and textual and cough based data sets. Latif et al. [28] reviewed data science research focusing on mitigation and diagnosis of COVID-19. The listed surveys mention few open source data sets and point towards the unavailability of open data resources challenging trustworthy and real-world operations of AI/ML-based techniques. Moreover, the critical analysis of possible AI innovations tackling COVID-19 have also highlighted open data as the first step towards the right direction [29, 30, 31]. Triggered by this challenge limiting the adoption of AI/ML-powered COVID-19 diagnosis, forecasting, and mitigation, we make the first effort in surveying research works based on open source data sets concerning COVID-19 pandemic.

The contributions of this article are manifold.

- We formulate a taxonomy of the research domain while identifying key characteristics of open source data sets in terms of their type, applications, and methods.
- We provide a comprehensive survey of the open-source COVID-19 data sets while categorizing them on data type, i.e., biomedical images, textual, and speech data. With each listed data set, we also describe the applied AI, big data, and statistical techniques.
- We provide a comparison of data sets in terms of their application, type, and size to provide valuable insights for data set selection.
- We highlight the future research directions and challenges for missing or limited data sets so that the research community can work towards the public availability of the data.

We are motivated by the fact that this survey will help researchers in the identification of appropriate open source data sets for their research. The comprehensive survey will also provide researchers with multiple directions to embark on an open data powered research against COVID-19.

Most of the articles included in this survey have not been rigorously peer-reviewed and published as pre-prints. However, their inclusion is necessary as the current pandemic situation requires rapid publishing process to propagate vital information on the pandemic. Moreover, the inclusion of non-peer-reviewed studies in this article is supported by their open source methods which can be independently verified. For the collection of the relevant literature review, we searched the online databases of Google scholar, BioRxiv, and medRxiv. The keywords employed were “COVID-19” and “data set”. We separately searched two online open source communities, i.e., Kaggle and Github for data sets that are not yet part of any publication. We focused on articles with applications of computer science and mathematics in general. We hope that our efforts will be fruitful in limiting the spread of COVID-19 through elaboration of open source scientific fact-finding efforts.

The rest of the article is organized as follows. In section 2, we detail the taxonomy of the research domain. Section 3 presents the comprehensive list of medical COVID-19 data sets divided into categories of CT scans and X-rays. Section 4 details a list of textual data sets classified into COVID-19 case report, cse report analysis, social media data, mobility data, NPI data, and scholarly article collections. Section 4.4 lists speech based data sets that diagnose COVID-19 from cough and breathing samples. In section 6, a comparison of listed data sets is provided in terms of openness, application, and data-type. Section 7 discusses the dimensions that need attention from scholars and future perspectives on COVID-19 open source research. Section 8 provides the concluding remarks for the article.

## 2 Taxonomy

Modern Information and Communication Technologies (ICT) help in combating COVID-19 on many fronts. These include research efforts towards:

- AI/ML based COVID-19 diagnosis and screening from medical images [32].
- COVID-19 case reports for transmission estimation and forecasting [3].
- COVID-19 emotional and sentiment analysis from social media [33].
- Semantic analysis of knowledge from the collection of scholarly articles covering COVID-19 [24].
- Application of AI enabled robots and drones to deliver food, medicine, and disinfect places [26].
- AI and ML based methods to find and evaluate drugs and medicines [34].
- Smart device based monitoring of lock-down and quarantine measures [35].
- Speech based breathing rate and stress detection [36].
- Cough based COVID-19 detection [37].

Some of these ICT powered techniques are data-driven. For example, detection of COVID-19 from medical images and cough samples requires samples from both non-COVID and COVID-19 positive cases. We focus on data-driven applications of ICT that are also open source.

We divide the data sets into three main categories, i.e., **(a)** medical images, **(b)** textual data, and **(c)** speech data. Medical images based data sets are mostly brought into service for screening and diagnosis of COVID-19. Medical images can belong to the class of CT scan, X-rays, MRT, or ultrasound. Most of the COVID-19 data sets contain either CT scans or X-rays [14]. Therefore, we categorize medical data sets into CT scans and X-ray classes. Moreover, some of the data sets consist of multiple types of images. Medical image-based diagnosis facilities are available at most hospitals and clinics. As COVID-19 test kits are short in supply and costly [38], medical image-based diagnosis lowers the burden on conventional PCR based screening. Medical image data sets are often pre-processed with segmentation and augmentation techniques. In medical images, segmentation leads to partitions of the image such that region of interest (infected region) is identified [39]. Image augmentation techniques include geometric transforms and kernel filters such that the size of the data set is enhanced. Consequently, the application of ML techniques that require bigger data sets is made possible avoiding overfitting [40]. The application of ML techniques on medical images has also led to the prediction of hospital stay duration in patients [41]. Medical image data sets should consider patient’s consent and preserve patient privacy [30].

Textual data sets serve four main purposes.

- Forecasting the visualizing and spread of COVID-19 based on reported cases.
- Analyzing public sentiment/opinion by tracking COVID-19 related posts on popular social media platforms.
- Collecting scholarly articles on COVID-19 for a centralized view on related research and application of information extraction/text mining.
- Studying the effect of mobility on COVID-19 transmissions.

The COVID-19 case reports can be further applied to: **(a)** compile data on regional levels (city, county, etc) [42], **(b)** visualize cases and forecast daily new cases, recoveries, etc [43], **(c)** study effects of mobility on number of new cases [44], and **(d)** study effect of non-pharmaceutical interventions (NPI) on COVID-19 cases [45, 46]. Speech data sets contain cough and breath sounds that can be employed to diagnose COVID-19 and its predict disease severity. ML, big data, and statistical techniques can be applied to the data sets for tasks related to prediction. Figure 1 illustrates a consolidated view of the taxonomy of COVID-19 open source data sets [13, 28].

**Figure 1:**
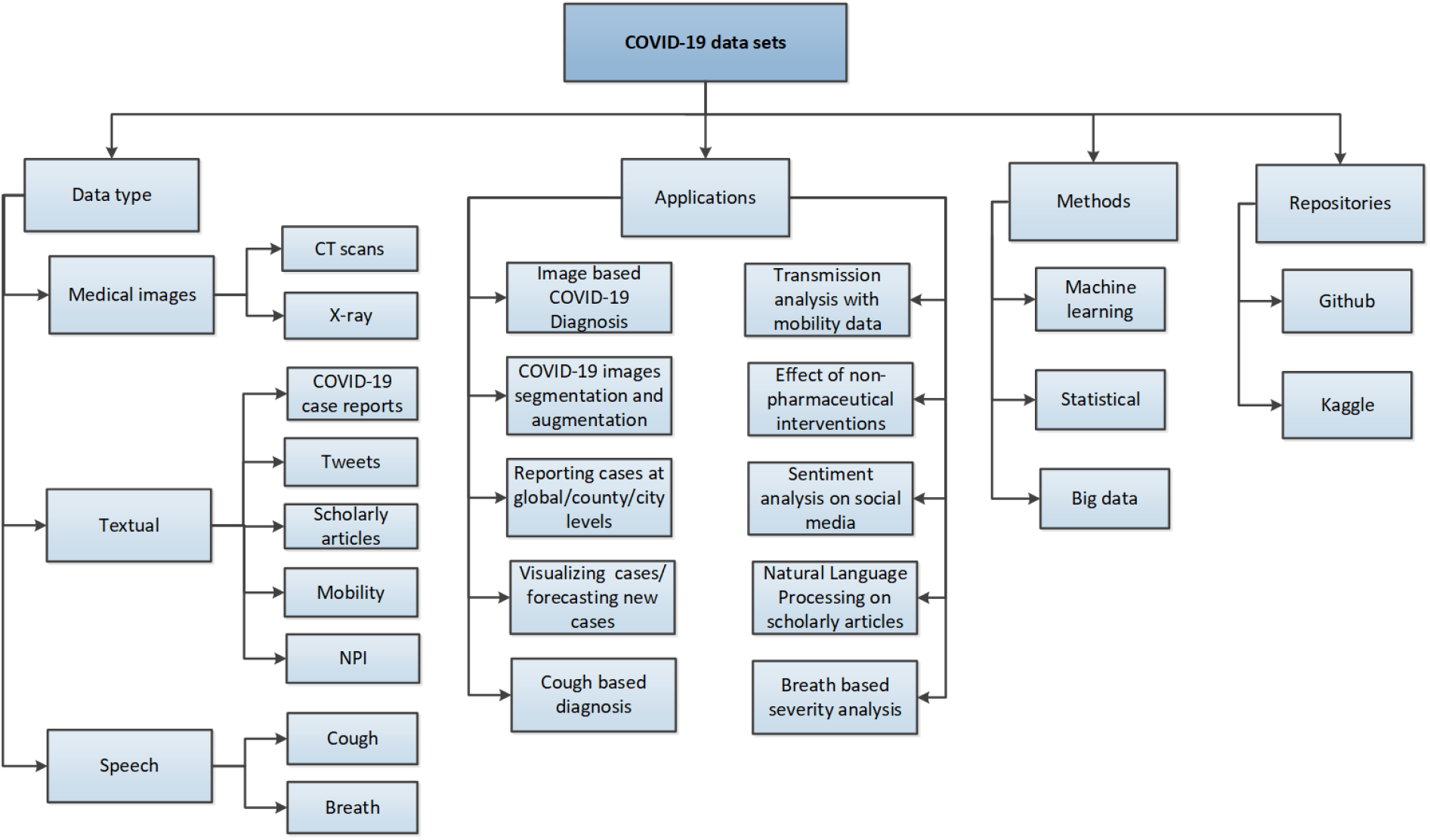
Taxonomy of COVID-19 open source data sets.

## 3 COVID-19 Medical image data sets

Medical images in the form of Chest CT scans and X-rays are essential for automated COVID-19 diagnosis. The AI powered COVID-19 diagnosis techniques can be as accurate as a human, save radiologist time, and perform diagnosis cheaper and faster than the common laboratory methods. We discuss CT scan and X-ray image data sets separately in the following subsections.

### 3.1 CT scans

Cohen et al. [23] describe the public COVID-19 image collection consisting of X-ray and CT-scans with ongoing updates. The data set consists of more than 125 images extracted from various online publications and websites. The data set specifically includes images of COVID-19 cases along with MERS, SARS, and ARDS based images. The authors enlist the application of deep and transfer learning on their extracted data set for identification of COVID-19 while utilizing motivation from earlier studies that learned the type of pneumonia from similar images [47]. Each image is accompanied by a set of 16 attributes such as patient ID, age, date, and location. The extraction of CT scan images from published articles rather than actual sources may lessen the image quality and affect the performance of the machine learning model. Some of the data sets available and listed below are obtained from secondary sources. The public data set published with this study is one of the pioneer efforts in COVID-19 image based diagnosis and most of the listed studies utilize this data set. The authors also listed perspective use cases and future directions for the data set [48].

Researcher [49] published a data set consisting of 275 CT scans of COVID-19 positive patients. The data set is extracted from 760 medRxiv and bioRxiv preprints about COVID-19. The authors also employed a deep convolutional network for training on the data set to learn COVID-19 cases for new data with an accuracy of around 85%. The model is trained on 183 COVID-19 positive CT scans and 146 negative cases. The model is tested on 35 COVID-19 positive CTs and 34 non-COVID CTs and achieves an F1 score of 0.85. Due to the small data set size, deep learning models tend to overfit. Therefore, the authors utilized transfer learning on the Chest X-ray data set released by NIH to fine-tune their deep learning model. The online repository is being regularly updated and currently consists of 349 CT images containing clinical findings of 216 patients.

Wang et al. [50] investigated a deep learning strategy for COVID-19 screening from CT scans. A total of 453 COVID-19 pathogen-confirmed CT scans were utilized along with typical viral pneumonia cases. The COVID-19 CT scans were obtained from various Chinese hospitals with an online repository maintained at ^3^. Transfer learning in the form of pre-trained CNN model (M-inception) was utilized for feature extraction. A combination of Decision tree and Adaboost were employed for classification with 83.9% accuracy.

Figure 2 depicts a generic work-flow of ML based COVID-19 diagnosis [51, 52]. The data set containing medical images is pre-processed with segmentation and augmentation techniques if necessary. Afterward, a ML pre-trained, fine-tuned, or built from scratch model is used to extract features for classification. The number of outputs classed is defined such as COVID-19 positive, negative, viral pneumonia. A classifier can classify each image from the data set based on its extracted feature [53].

**Figure 2:**
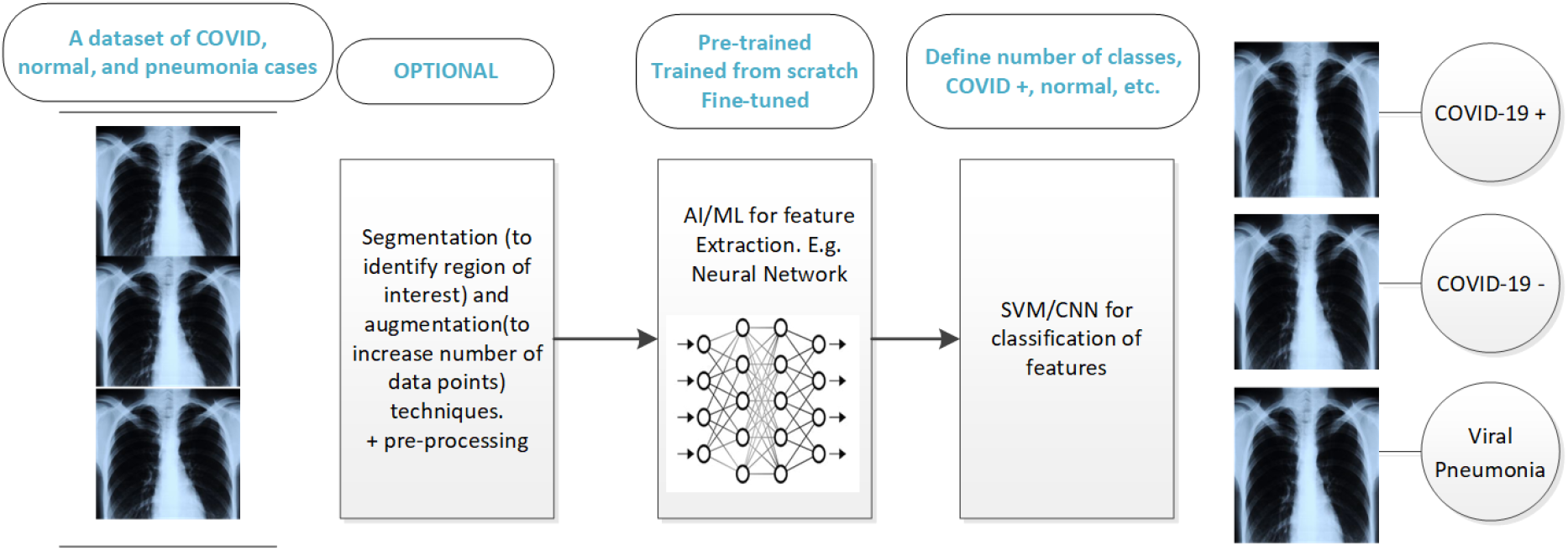
A generic work-flow of AI/ML based COVID-19 diagnosis.

#### 3.1.1 Segmentation data sets

Segmentation helps health service providers to quickly and objectively evaluate the radiological images. Segmentation is a pre-processing step that outlines the region of interest, e.g., infected regions or lesions for further evaluation. Shan et al. [54] obtained CT scan images from COVID-19 cases based mostly in Shanghai for deep learning-based lung infection quantification and segmentation. However, their data set is not public. The deep learning-based segmentation utilizes VB-Net, a modified 3-D convolutional neural network (CNN), to segment COVID-19 infection regions in CT scans. The proposed system performs auto-contouring of infection regions, accurately estimates their shapes, volumes, and percentage of infection. The system is trained using 249 COVID-19 patients and validated using 300 new COVID-19 patients. Radiologists contributed as a human in the loop to iteratively add segmented images to the training data set. Two radiologists contoured the infectious regions to quantitatively evaluate the accuracy of the segmentation technique. The proposed system and manual segmentation resulted in 90% dice similarity coefficients.

Other than the published articles, few online efforts have been made for image segmentation of COVID-19 cases. A COVID-19 CT lung and infection segmentation data set is listed as open source [55, 56]. The data set consists of 20 COVID-19 CT scans labeled into left, right, and infectious regions by two experienced radiologists and verified by another radiologist ^4^. Three segmentation benchmark tasks have also been created based on the data set ^5^.

Another such online initiative is the COVID-19 CT segmentation data set ^6^. The segmented data set is hosted by two radiologists based in Oslo. They obtained images from a repository hosted by the Italian society of medical and interventional radiology (SIRM) ^7^. The obtained images were segmented by the radiologist using 3 labels, i.e., ground-glass, consolidation, and pleural effusion. As a result, a data set that contains 100 axial CT slices from 60 patients with manual segmentation in the form of JPG images is formed. Moreover, the radiologists also trained a 2D multi-label U-Net model for automated semantic segmentation of images.

#### 3.1.2 Online data sets

In this subsection, we list COVID-19 data set initiatives that are public but are not associated with any publication.

The Coronacases Initiative shares 3D CT scans of confirmed cases of COVID-19 ^8^. Currently, the web repository contains 3D CT images of 10 confirmed COVID-19 cases shared for scientific purposes. The British Society of Thoracic Imaging (BSTI) in collaboration with Cimar UK’s Imaging Cloud Technology deployed a free to use encrypted and anonymised online portal to upload and download medical images related to COVID-19 positive and suspected patients ^9^. The uploaded images are sent to a group of BSTI experts for diagnosis. Each reported case includes data regarding the age, sex, PCR status, and indications of the patient. The aim of the online repository is to provide COVID-19 medical images for reference and teaching. The SIRM is hosting radiographical images of COVID-19 cases ^10^. Their data set has been utilized by some of the cited works in this article. Another open source repository for COVID-19 radiographic images is Radiopaedia ^11^. Multiple studies [57, 58] employed this data set for their research.

### 3.2 X-ray

Researchers [59] present COVID-Net, a deep convolutional network for COVID-19 diagnosis based on Chest X-ray images. Motivated by earlier efforts on radiography based diagnosis of COVID-19, the authors make their data set and code accessible for further extension. The data set consists of 13,800 chest radiography images from 13,725 patient cases from three open access data repositories. The COVID-Net architecture consists of two stages. In the first stage, residual architecture design principles are employed to construct a prototype neural network architecture to predict either of **(a)** normal, **(b)** non-COVID infection, and **(c)** COVID-19 infections. In the second stage, the initial network design prototype, data, and human-specific design requirements act as a guide to a design exploration strategy to learn the parameters of deep neural network architecture. The authors also audit the COVID-Net with the aim of transparency via examination of critical factors leveraged by COVID-Net in making detection decisions. The audit is executed with GSInquire which is a commonly used AI/ML explainability method [60].

Author in [61] utilized the data set of Cohen et al. [23] and proposed COVIDX-Net, a deep learning framework for automatic COVID-19 detection from X-ray images. Seven different deep CNN architecture, namely, VGG19, DenseNet201, InceptionV3, ResNetV2, InceptionResNetV2, Xception, and MobileNetV2 were utilized for performance evaluation. The VGG19 and DenseNet201 model outperform other deep neural classifiers in terms of accuracy. However, these classifiers also demonstrate higher training times.

Apostolopoulos et al [62] merged the data set of Cohen et al. [23], a data set from Kaggle ^12^, and a data set of common bacterial-pneumonia X-ray scans [63] to train a CNN to distinguish COVID-19 from common pneumonia. Five CNNs namely VGG19, MobileNet v2, Inception, Xception, and Inception ResNet v2 with common hyper-parameters were employed. Results demonstrate that VGG19 and MobileNet v2 perform better than other CNNs in terms of accuracy, sensitivity, and specificity.

The researchers extended their work in [58] to extract bio-markers from X-ray images using a deep learning approach. The authors employ MobileNetv2, a CNN is trained for the classification task for six most common pulmonary diseases. MobileNetv2 extracts features from X-ray images in three different settings, i.e., from scratch, with the help of transfer learning (pre-trained), and hybrid feature extraction via fine-tuning. A layer of Global Average Pooling was added over MobileNetv2 to reduce overfitting. The extracted features are input to a 2500 node neural network for classification. The data set include recent COVID-19 cases and X-rays corresponding to common pulmonary diseases. The COVID-19 images (455) are obtained from Cohen et al. [23], SIRM, RSNA, and Radiopaedia. The data set of common pulmonary diseases is extracted from a recent study [63] among other sources. The training from scratch strategy outperforms transfer learning with higher accuracy and sensitivity. The aim of the research is to limit exposure of medical experts with infected patients with automated COVID-19 diagnosis.

Researchers [64] merged the data set of Cohen et al. [23] (50 images) and a data set from Kaggle ^13^ (50 images) for application of three pre-trained CNNs, namely, ResNet50, InceptionV3, and InceptionResNetV2 to detect COVID-19 cases from X-ray radiographs. The data set was equally divided into 50 normal and 50 COVID-19 positive cases. Due to the limited data set, deep transfer learning is applied that requires smaller data set to learn and classify features. The ResNet50 provided the highest accuracy for classifying COVID-19 cases among the evaluated models.

Authors in [51] propose Support Vector Machine (SVM) based classification of X-ray images instead of predominately employed deep learning models. The authors argue that deep learning models require large data sets for training that are not available currently for COVID-19 cases. The data set brought to service in this article is an amalgam of Cohen et al. [47], a data set of Kaggle ^14^, and data set of Kermany et. [63]. Author of [62] also utilized data from same sources. The data set consists of 127 COVID-19 cases, 127 pneumonia cases, and 127 healthy cases. The methodology classifies the X-ray images into COVID-19, pneumonia, and normal cases. Pre-trained networks such as AlexNet, VGG16, VGG19, GoogleNet, ResNet18, ResNet50, ResNet101, InceptionV3, InceptionResNetV2, DenseNet201, XceptionNet, MobileNetV2 and ShuffleNet are employed on this data set for deep feature extraction. The deep features obtained from these networks are fed to the SVM classifier. The accuracy and sensitivity of ResNet50 plus SVM is found to be highest among CNN models.

Similar to Sethy et al. [51], Afshar et al. [32] also negated the applicability of DNNs on small COVID-19 data sets. The authors proposed a capsule network model (COVID-CAPS) for the diagnosis of COVID-19 based on X-ray images. Each layer of a COVID-CAPS consists of several Capsules, each of which represents a specific image instance at a specific location with the help of several neurons. The length of a Capsule determines the existence probability of the associated instance. COVID-CAPS uses four convolutional and three capsule layers and was pre-trained with transfer learning on the public NIH data set of X-rays images for common thorax diseases. COVID-CAPS provides a binary output of either positive or negative COVID-19 case. The COVID-CAPS achieved an accuracy of 95.7%, a sensitivity of 90%, and specificity of 95.8%.

Many authors have applied augmentation techniques on COVID-19 image data sets. Authors [52] utilized data augmentation techniques to increase the number of data points for CNN based classification of COVID-19 X-ray images. The proposed methodology adds data augmentation to basic steps of feature extraction and classification. The authors utilize the data set of Cohen et al. [23]. The authors design five deep learning model for feature extraction and classification, namely, custom-made CNNs trained from scratch, transfer learning-based fine-tuned CNNs, proposed novel COVID-RENet, dynamic feature extraction through CNN and classification using SVM, and concatenation of dynamic feature spaces (COVID-RENet and VGG-16 features) and classification using SVM. SVM classification is brought to serve to further increase the accuracy of the task. The results showed that the proposed COVID-RENet and Custom VGG-16 models accompanied by the SVM classifier show better performance with approximately 98.3% accuracy in identifying COVID-19 cases.

Researchers [65] formulated a data set of COVID-19 and non-COVID-19 cases containing both X-ray images and CT scans. Augmentation techniques are applied on the data set to obtain approximately 17000 X-ray and CT images. The data set is divided into main categories of X-ray and CT images. The X-ray images comprise of 4044 COVID-19 positive and 5500 non-COVID images. The CT-scan images comprise of 5427 COVID-19 positive and 2628 non-COVID images. These works on augmentation of COVID-19 images resolve the issue of data scarcity for deep learning techniques. However, further investigation is required to determine the effectiveness of ML techniques in detecting COVID-19 cases from augmented data sets.

Authors [66] contributed towards a single COVID-19 X-ray image database for AI applications based on four sources. The aim of the research was to explore the possibility of AI application for COVID-19 diagnosis. The source databases were Cohen et al. [23], Italian Society of Medical and Interventional Radiology data set, images from recently published articles, and a data set hosted at Kaggle ^15^. The cumulative data set contains 190 COVID-19 images, 1345 viral pneumonia images, and 1341 normal chest x-ray images. The authors further created 2500 augmented images from each category for the training and validation of four CNNs. The four tested CNNs are AlexNet, ResNet18, DenseNet201, and SqueezeNet for classification of Xray images into normal, COVID-19, and viral pneumonia cases. The SqueezeNet outperformed other CNNs with 98.3% accuracy and 96.7% sensitivity. The collective database can be found at ^16^.

Born et al. [67] advocated the role of ultra-sound images for COVID-19 detection. Compared to CT scans, ultra-sound is a non-invasive, cheap, and portable medical imaging technique. First, the authors aggregated data in an open source repository ^17^ named Point-of-care Ultrasound (POCUS). The data set consists of 1103 images (654 COVID-19, 277 bacterial pneumonia, and 172 healthy controls) extracted from 64 online videos and published research works. The main sources of the data were grepmed.com, thepocusatlas.com, butterflynetwork.com, and radiopaedia.org. Data augmentation techniques are also used to diversify the data. Afterward, the authors train a deep CNN (VGG-16) named POCOVID-Net on the three-class data set to achieve an accuracy of 89% and sensitivity for COVID of 96%. Lastly, the authors provide an open access medical service named POCOVIDScreen to classify and predict lung ultra-sound images ^18^.

A comprehensive list of AI-based COVID-19 research can be found at [68]. A list open source data sets on the Kaggle can be found at ^19^. A crowd-sourced list of open access COVID-19 projects can be found at ^20^.

## 4 COVID-19 Textual data sets

COVID-19 case reports, global and county-level dashboards, case report analysis, mobility data, social media posts, NPI, and scholarly article collections are detailed in the following subsections.

## 4.1 COVID-19 case reports

The earliest and most noteworthy data set depicting the COVID-19 pandemic at a global scale was contributed by John Hopkins University [43]. The authors developed an online real-time interactive dashboard first made public in January 2020 ^21^. The dashboard lists the number of cases, deaths, and recoveries divided into country/provincial regions. A data is more detailed to the city level for the USA, Canada, and Australia. A corresponding Github repository of the data is also available ^22^ and Datahub repository is available at ^23^. The data collection is semi-automated with main sources are DXY (a medical community ^24^) and WHO. The DXY community collects data from multiple sources and updated every 15 minutes. The data is regularly validated from multiple online sources and health departments. The aim of the dashboard was to provide the public, health authorities, and researchers with a user-friendly tool to track, analyze, and model the spread of COVID-19. Authors [69] employed four supervised ML models including SVM and linear regression on this data set the predict the number of new cases, deaths, and recoveries.

Dey et al. [70] analyzed the epidemiological outbreak of COVID-19 using a visual exploratory data analysis approach. The authors utilized publicly available data sets from WHO, the Chinese Center for Disease Control and Prevention, and Johns Hopkins University for cases between 22 January 2020 to 16 February 2020 all around the globe. The data set consisted of time-series information regarding the number of cases, origin country, recovered cases, etc. The main objective of the study is to provide time-series visual data analysis for the understandable outcome of the COVID-19 outbreak.

Liu et al. [71] formulated a spatio-temporal data set of COVID-19 cases in China on the daily and city levels. As the published health reports are in the Chinese language, the authors aim to facilitate researchers around the globe with data set translated to English. The data set also divides the cases to city/county level for analysis of city-wide pandemic spread contrary to other data sets that provide county/province level categorizations ^25^. The data set consists of essential statistics for academic research, such as daily new infections, accumulated infections, daily new recoveries, accumulated recoveries, daily new deaths, etc. Each of these statistics is compiled into a separate CSV file and made available on Github. The first two authors did cross-validation of their data extraction tasks to reduce the error rate.

Researchers [19, 72] list and maintain the epidemiological data of COVID-19 cases in China. The data set contains individual-level information of laboratory-confirmed cases obtained from city and provincial disease control centers. The information includes **(a)** key dates including the date of onset of disease, date of hospital admission, date of confirmation of infection, and dates of travel, **(b)** demographic information about the age and sex of cases, **(c)** geographic information, at the highest resolution available down to the district level, **(d)** symptoms, and **(e)** any additional information such as exposure to the Huanan seafood market. The data set is updated regularly. The aim of the open access line list data is to guide the public health decision-making process in the context of the COVID-19 pandemic.

Killeen et al. [42] accounted for the county-level data set of COVID-19 in the US. The machine-readable data set contains more than 300 socioeconomic parameters that summarize population estimates, demographics, ethnicity, education, employment, and income among other healthcare system-related metrics. The data is obtained from the government, news, and academic sources. The authors obtain time-series data from [43] and augment it with activity data obtained from SafeGraph. Details of the SafeGraph data set can be found in the Section 4.5. A collection of country specific case reports and articles can be found at Harvard Dataverse Repository ^26^.

### 4.2 COVID-19 case report analysis

Based on the open source data sets, we list various open source analysis efforts in this sub-section. Most of listed research works in the below sections are accompanied by both open source data and code. The COVID-19 textual data analysis serve varying purposes, such as, forecasting COVID-19 transmission from China, estimating the effect of NPI and mobility on number of cases, estimating the serial interval and reproduction rate.

#### 4.2.1 COVID-19 transmission analysis

Kucharski et al. [4] modeled COVID-19 cases based on data set of cases from and international cases that originated from Wuhan. The purpose of the study was to estimate human-to-human transmissions and virus outbreaks if the virus was introduced in a new region. The four time-series data sets used were: the daily number of new internationally exported cases, the daily number of new cases in Wuhan with no market exposure, the daily number of new cases in China, and the proportion of infected passengers on evacuation flights between December 2019 and February 2020. The study while employing stochastic modeling found that the R*0* declined from 2.35 to 1.05 after travel restrictions were imposed in Wuhan. The study also found that if four cases are reported in a new area, there is a 50% chance that the virus will establish within the community.

Researchers [73] described an econometric model to forecast the spread and prevalence of COVID-19. The analysis is aimed to aid public health authorities to make provisions ahead of time based on the forecast. A time-series database was built based on statistics from Johns Hopkins University and made public in CSV format. Auto-Regressive Integrated Moving Average (ARIMA) model prediction on the data to predict the epidemiological trend of the prevalence and incidence of COVID-2019. The ARIMA model consists of an autoregressive model, moving average model, and seasonal autoregressive integrated moving average model. The ARIMA model parameters were estimated by autocorrelation function. ARIMA (1,0,4) model was selected for the prevalence of COVID-2019 while ARIMA (1,0,3) was selected as the best ARIMA model for determining the incidence of COVID-19. The research predicted that if the virus does not develop any new mutations, the curve will flatten in the near future.

Researcher [74] utilize reported death rates in South Korea in combination with population demographics for correction of under-reported COVID-19 cases in other countries. South Korea is selected as a benchmark due to its high testing capacity and well-documented cases. The author correlates the under-reported cases with limited sampling capabilities and bias in systematic death rate estimation. The author brings to service two data sets. One of the data sets is WHO statistics of daily country-wise COVID-19 reports. The second data set is demographic database maintained by the UN. This data set is limited from 2007 onwards and hosted on Kaggle for country wise analysis ^27^. The adjustment in number of COVID-19 cases is achieved while comparing two countries and computing their Vulnerability Factor which is based on population ages and corresponding death rates. As a result, the Vulnerability Factor of countries with higher age population is greater than one leading to higher death rate estimations. A complete work-flow of the analysis is also hosted on Kaggle ^28^.

#### 4.2.2 COVID-19 NPI analysis

Kraemer et al. [44] analyzed the effect of human mobility and travel restrictions on spread of COVID-19 in China. Real-time and historical mobility data from Wuhan and epidemiological data from each province were employed for the study (source: Baidu Inc.). The authors also maintain a list of cases in Hubei and a list of cases outside Hubei. The data and code can be found at ^29^. The study found that before the implementation of travel restrictions, the spatial distribution of COVID-19 can be highly correlated to mobility. However, the correlation is lower after the imposition of travel restrictions. Moreover, the study also estimated that the late imposition of travel restrictions after the onset of the virus in most of the provinces would have lead to higher local transmissions. The study also estimated the mean incubation period to identify a time frame for evaluating early shifts in COVID-19 transmissions. The incubation period was estimated to be 5.1 days.

Researchers [75] provide another study for evaluating the effects of travel restrictions on COVID-19 transmissions. The authors quantify the impact of travel restrictions in early 2020 with respect to COVID-19 cases reported outside China using statistical analysis. The authors obtained an epidemiological data set of confirmed COVID-19 cases from government sources and websites. All confirmed cases were screened using RT-PCR. The quantification of COVID-19 transmission with respect to travel restrictions was carried out for the number of exported cases, the probability of a major epidemic, and the time delay to a major epidemic.

Lai et al. [76] quantitatively studied the effect of NPI, i.e., travel bans, contact reductions, and social distancing on the COVID-19 outbreak in China. The authors modeled the travel network as Susceptible-exposed-infectious-removed (SEIR) model to simulate the outbreak across cities in China in a proposed model named Basic Epidemic, Activity, and Response COVID-19 model. The authors used epidemiological data in the early stage of the epidemic before the implementation of travel restrictions. This data was used to determine the effect of NPI on onset delay in other regions with first case reports as an indication. The authors also obtained large scale mobility data from Baidu location-based services which report 7 billion positioning requests per day. Another historical data set from Baidu was obtained for daily travel patterns during the Chinese new year celebrations which coincided with the COVID-19 outbreak. The study estimated that there were approximately 0.1 Million COVID-19 cases in China as of 29 February 2020. Without the implementation of NPI, the cases were estimated to increase 67 fold. The impact of various restrictions was varied with early detection and isolation preventing more cases than the travel restrictions. In the case of a three-week early implementation of NPI, the cases would have been 95% less. On the contrary, if the NPI were implemented after a further delay of 3 weeks, the COVID-19 cases would have increased 18 times.

A study on a similar objective of investigating the impact of NPI in European countries was carried out in [45]. At the start of pandemic spread in European countries, NPI were implemented in the form of social distancing, banning mass gathering, and closure of educational institutes. The authors utilized a semi-mechanistic Bayesian hierarchical model to evaluate the impact of these measures in 11 European countries. The model assumes that any change in the reproductive number is the effect of NPI. The model also assumed that the reproduction number behaved similarly across all countries to leverage more data across the continent. The study estimates that the NPI have averted 59000 deaths up till 31 March 2020 in the 11 countries. The proportion of the population infected by COVID-19 is found to be highest in Spain followed by Italy. The study also estimated that due to mild and asymptomatic infections many fold low cases have been reported and around 15% of Spain population was infected in actual with a mean infection rate of 4.9%. The mean reproduction number was estimated to be 3.87. Real-time data was collected from ECDC (European Centre of Disease Control) for the study ^30^.

Wells et al. [77] studied the impact of international travel and border control restrictions on COVID-19 spread. The research work utilized daily case reports from December 8, 2019 to February 15, 2020 in China and county-wise airport connectivity with China to estimate the risk of COVID-19 transmissions. A total of 63 countries have direct flight connectivity with mainland China. The COVID-19 transmission/importation risk was assumed to be proportional to the number of airports with direct flights from China. It was estimated that an average reduction of 81.2% exportation rate occurred due to the travel and lockdown restrictions. Health questionnaire regarding exposure at least a week prior to arrival is estimated to identify 95% of the cases during incubation period. It is also estimated that if a case is identified via contact tracing within 5 days of exposure, the chances of its travel during the incubation period are reduced by 24.7%.

Researcher [78] investigated the transmission control measures of COVID-19 in China. The authors compiled and analyzed a unique data set consisting of case reports, mobility patterns, and public health intervention. The COVID-19 case data were collected from official reports of the health commission. The mobility data were collected from location-based services employed by Social media applications such as WeChat. The travel pattern from Wuhan during the spring festival was constructed from Baidu migration index ^31^. The study found that the number of cases in other provinces after the shutdown of Wuhan can be strongly related to travelers from Wuhan. In cities with a lesser population, the Wuhan travel ban resulted in a delayed arrival (+2.91 days) of the virus. Cities that implemented the highest level emergency before the arrival of any case reported 33.3% lesser number of cases. The low level of peak incidences per capita in provinces other than Wuhan also indicates the effectiveness of early travel bans and other emergency measures. The study also estimated that without the Wuhan travel band and emergency measures, the number of COVID-19 cases outside Wuhan would have been around 740000 on the 50th day of the pandemic. In summary, the study found a strong association between the emergency measures introduced during spring holidays and the delay in epidemic growth of the virus.

#### 4.2.3 COVID-19 reproduction rate analysis

Tindale et al. [79] study the COVID-19 outbreak to estimate the incubation period and serial interval distribution based on data obtained in Singapore (93 cases) and Tianjin (135). The incubation period is the period between exposure to an infection and the appearance of the first symptoms. The data was made available to the respective health departments. The serial interval can be used to estimate the reproduction number (R*0*) of the virus. Moreover, both serial interval and incubation period can help identify the extent of pre-symptomatic transmissions. With more than a months data of COVID-19 cases from both cities, The mean serial interval was found to be 4.56 days for Singapore and 4.22 days for Tianjin. The mean incubation period was found to be 7.1 days for Singapore and 9 days for Tianjin.

Researchers [80] investigated the serial interval of COVID-19 based on publicly reported cases. A total of 468 COVID-19 transmission events reported in China outside of Hubei Province between January 21, 2020, and February 8, 2020 formulated the data set. The data is compiled from reports of provincial disease control centers. The data indicated that in 59 of the cases, the infectee developed symptoms earlier than the infector indicated pre-symptomatic transmission. The mean serial interval is estimated to be 3.96 with a standard deviation of 4.75. The mean serial interval of COVID-19 is found to be lower than similar viruses of MERS and SARS. The production rate (R*0*) of the data set is found to be 1.32.

Author in [81] presented a framework for serial interval estimation of COVID-19. As the virus is easily transmitted in a community from an infected person, it is important to know the onset of illness in primary to secondary transmissions. The date of illness onset is defined as the date on which a symptom relevant to COVID-19 infection appears. The serial interval refers to the time between successive cases in a chain of disease transmission. The authors obtain 28 cases of pairs of infector-infectee cases published in research articles and investigation reports and rank them for credibility. A subset of 18 high credible cases are selected to analyze that the estimated median serial interval lies at 4.0 days. The median serial interval of COVID-19 is found to be smaller than SARS. Moreover, it is implied that contact tracing methods may not be effective due to the rapid serial interval of infector-infectee transmissions.

### 4.3 Social media data

Researchers [82] contributed towards a publicly available ground truth textual data set to analyze human emotions and worries regarding COVID-19. The initial findings were termed as Real World Worry Dataset (RWWD). In the current global crisis and lock-downs, it is very essential to understand emotional responses on a large scale. The authors requested multiple participants from UK on 6th and 7th April (Lock-down, PM in ICU) to report their emotions and formed a data set of 5000 texts (2500 short and 2500 long texts). The number of participants was 2500. Each participant was required to provide a short tweet-sized text (max 240 characters) and a long open-ended text (min 500 characters). The participants were also asked to report their feelings about COVID-19 situations using 9-point scales (1 = not at all, 5 = moderately, 9 = very much). Each participant rated how worried they were about the COVID-19 situation and how much anger, anxiety, desire, disgust, fear, happiness, relaxation, and sadness they felt. One of the emotions that best represented their emotions was also selected. The study found that anxiety and worry were the dominant emotions. STM package from R was reported for topic modeling. The most prevalent topic in long texts related to the rules of lock-down and the second most prevalent topic related to employment and economy. In short texts, the most prominent topic was government slogans for lock-down.

A large-scale Twitter stream API data set for scientific research into social dynamic of COVID-19 was presented in [83]. The data set is maintained by the Panacea Lab at Georgia State University with dedicated efforts starting on March 11, 2020. The data set consists of more than 424 million tweets in the latest version with daily updates. The data set consists of tweets in all languages with prevalence of English, Spanish, and French. Several keywords, such as, COVID19, CoronavirusPandemic, COVID-19, 2019nCoV, CoronaOutbreak, and coronavirus were used to filter results. The data set consists of two TSV files, i.e., a full data set and one that has been cleaned with no re-tweets. The data set also contains separate CSV files indicating top 1000 frequent terms, top 1000 bigrams, and top 1000 trigrams.

Chen et al. [84] describe a multilingual coronavirus data set with the aim of studying online conversation dynamics. The social distancing measure has resulted in abrupt changes in the society with the public accessing social platforms for information and updates. Such data sets can help identify rumors, misinformation, and panic among the public along with other sentiments from social media platforms. Using Twitter’s streaming API and Tweepy, the authors began collecting tweets from January 28, 2020 while adding keywords and trending accounts incrementally in the search process. At the time of publishing, the data set consisted of over 50 million tweets and 450GB of raw data.

Authors in [85] collected a Twitter data set of Arabic language tweets on COVID-19. The aim of the data set collection is to study the pandemic from a social perspective, analyze human behavior, and information spread with special consideration to Arabic speaking countries. The data set collection was started in March 2020 using twitter API and consists of more than 2,433,660 Arabic language tweets with regular additions. Arabic keywords were used to search for relevant tweets. Hydrator and TWARC tools are employed for retrieving the full object of the tweet. The data set stores multiple attributes of a tweet object including the ID of the tweet, username, hashtags, and geolocation of the tweet.

Yu et al. [86] compiled a data set from Twitter API solely based on the institutional and news channel tweets based on multiple countries including US, UK, China, Spain, France, and Germany. A total of 69 Twitter accounts were followed with 17 from government and international organizations including WHO, EU commission, CDC, ECDC. 52 news media outlets were monitored including NY Times, CNN, Washington Post, and WSJ.

Researcher [5] analyzes a data set of tweets about COVID-19 to explore the policies and perceptions about the pandemic. The main objective of the study is to identify public response to the pandemic and how the response varies time, countries, and policies. The secondary objective is to analyze the information and misinformation about the pandemic is presented and transmitted. The data set is collected using Twitter API and covers 22 January to 13 March 2020. The corpus contains 6,468,526 tweets based on different keywords related to the virus in multiple languages. The data set is being continuously updated. The authors propose the application of Natural Language Processing, Text Mining, and Network Analysis on the data set as their future work. Similar data sets of Twitter posts regarding COVID-19 can be found at Github ^32^ and Kaggle ^33^.

Zarei et al. [87] gather social media content from Instagram using hashtags related to COVID-19 (coronavirus, covid19, and corona etc.). The authors found that 58% of the social media posts concerning COVID-19 were in English Language. The authors proposed the application of fake new identification and social behavior analysis on their data set.

Sarker et al. [88] mined Twitter to analyze symptoms of COVID-19 from self-reported users. The authors identified 203 COVID-19 patients while searching Twitter streaming API with expressions related to self-report of COVID-19. The patients reported 932 different symptoms with 598 unique lexicons. The most frequently reported COVID-19 symptoms were fever (65%) and cough (56%). The reported symptoms were compared with clinical findings on COVID-19. It was found that anosmia (26%) and ageusia (24%) reported on Twitter were not found in the clinical studies. A generic workflow of ML and NLP application on social media data is illustrated in figure 3.

**Figure 3:**
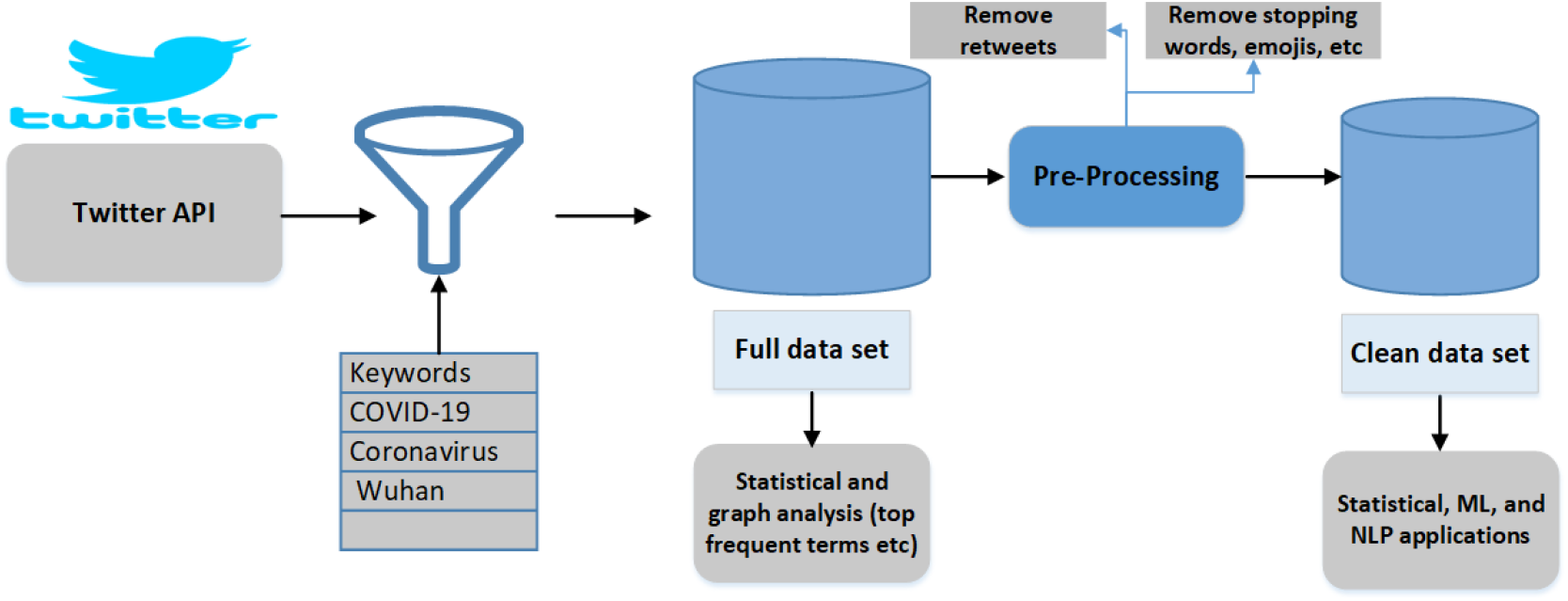
A generic work-flow of Social media based ML and NLP applications [83].

### 4.4 Scholarly articles

Several articles have performed bibliometric research on scientific works focused on COVID-19. The Allen Institute for AI with other collaborators started an initiative for collecting articles on COVID-19 research named CORD-19 [6]. The data set was initiated with 28K articles now contains more than 52k articles and 41k full texts. Multiple repositories such as PMC, BioRxiv, MedRxiv, and WHO were searched with queries related to COVID-19 (“COVID-19”, “Coronavirus”, “Corona virus”, “2019-nCoV”, etc.). Along with the article’s data set, a metadata file is also provided which includes each article DOI and publisher among other information. The data set is also divided into commercial and non-commercial subsets. The duplicate articles were clustered based on publication ID/DOI and filtered to remove duplication. Design challenges such as machine-readable text, copyright restrictions, and clean canonical meta-data were considered while collecting data. The aim of the data set collection is to facilitate information retrieval, extraction, knowledge-based discovery, and text mining efforts focused on COVID-19 management policies and effective treatment. The data set has been popular among the research community with more than 1.5 million views and more than 75k downloads. A competition at Kaggle based on information retrieval from the proposed data set is also active. On the other hand, several publishers have created separate sections for COVID-19 research and listed on their website. Ahamed et al. [89] applied graph-based techniques on this data set to study three topic related to COVID-19. Researchers [90] applied Association Rule Text Mining (ARTM) and information cartography techniques on the same data set. ARTM highlights distinguished terms and the association between them after parsing text documents while information cartography extracts structured knowledge from association rules.

Researchers [91] provided a scoping review of 65 research articles published before 31 January 2020 indicating early studies on COVID-19. The review followed a five-step methodological framework for the scoping review as proposed in [92]. The authors searched multiple online databases including bioRxiv, medRxiv, Google scholar, PubMed, CNKI, and WanFang Data. The searched terms included “nCoV”, “2019 novel coronavirus”, and “2019-nCoV” among others. The study found that approximately 90% of the published articles were in the English language. The largest proportion (38.5%) of articles studied the causes of COVID-19. Chinese authors contributed to most of the work (67.7%). The study also found evidence of virus origin from the Wuhan seafood market. However, specific animal association of the COVID-19 was not found. The most commonly reported symptoms were fever, cough, fatigue, pneumonia, and headache form the studies conduction clinical trails of COVID-19. The surveyed studies have reported masks, hygiene practices, social distancing, contact tracing, and quarantines as possible methods to reduce virus transmission. The article sources are available as supplementary resources with the article.

Researchers [24] detailed a systematic review and critical appraisal of prediction models for COVID-19 diagnosis and prognosis. Multiple publication repositories such as PubMed were searcher for articles that developed and validated COVID-19 prediction models. Out of the 2696 available titles and 27 studies describing 31 prediction models were included for the review. Three of these studies predicted hospital admission from pneumonia cases. Eighteen studies listed COVID-19 diagnostic models out of which 13 were ML-based. Ten studies detailed prognostic models that predicted mortality rates among other parameters. The critical analysis utilized PROBAST, a tool for risk and bias assessment in prediction models [93]. The analysis found that the studies were at high risk of bias due to poorly reported models. The study recommended that COVID-19 diagnosis and prognosis models should adhere to transparent and open source reporting methods to reduce bias and encourage realtime application.

Researcher from Berkeley lab have developed a web search portal for data set of scholarly articles on COVID-19 ^34^. The data set is composed of several scholarly data sets including Wang et al. [6], LitCovid, and Elsevier Novel Corona virus Information Center. The continuously expanding data set contains approximately 60K articles with 16K specifically related to COVID-19. The search portal employs NLP to look for related articles on COVID-19 and also provides valuable insights regarding the semantic of the articles.

### 4.5 Mobility data sets

Mobility data sets during the COVID-19 pandemic are essential to establish a relation between the number of cases (transmitted) and mobility patterns and observe the global response of communities in NPI restrictions. There are a number of open source mobility data sets providing information with varying features. Mobility data sets can be investigated to answer questions like what is the effect of COVID-19 on travel? Did people stay at home during the lock-down? Is their a correlation between high death rates and high mobility?

A global mobility data set of more than 150 countries collected from Google location services can be found at Google ^35^. It presents reports available in PDF format with a breakdown of countries and regions. The reports include a summary of changes in retail, recreation, supermarket, pharmacy, park, public transport, workplace, and residential visits. The privacy of users is ensured with aggregated and anonymised data that contains no identifiable personal information. A summary of the Google mobility reports in CSV format can be found at Kaggle ^36^.

Apple made a similar data set available on mobility based on user requests to location services across the globe ^37^. User privacy was addressed with anonymised records as data sent from devices is associated with random rotating identifiers. The data set available in CSV format compared the mobility with a baseline set on 13th January 2020. The data set contains information on the country/region, sub-region, or city level. The GeoDS lab at the University of Wisconsin-Madison has developed a web application identifying mobility patterns across the U.S ^38^. The data set is based on reports from SafeGraph ^39^ and Descartes Labs ^40^. The baseline is formed between the two weekends occurring before the lock-down measures were announced in the U.S. The data set provides fine-grained details up to county levels.

SafeGraph is a digital footprint platform that aggregates location-based data from multiple applications in the U.S. The journalistic data is used to infer the implementation of lock-down measures at the county level. The data set is envisioned to serve the scientific community in general and ML applications specifically for epidemiological modeling. Some of the research works listed in the above sections have utilized mobility data from Baidu location services ^41^. Limited data about commercial flights can be obtained from the Flirt tool ^42^.

### 4.6 NPI data sets

NPI is the collection of wide range of measures adopted by governments to curb the COVID-19 pandemic. NPI data sets are essential to the study of COVID-19 transmissions and analyze the effect of NPI on COVID-19 cases (infections, deaths, etc) for better policy and decisions at government level [46].

A team of academics and students at Oxford University systematically collected publicly available data from every part of the world into a Stringency Index [94]. The Stringency Index consists of information on government policies and a score to indicate their stringency. A total of 17 indicators are listed grouped into four classes of containment and closure (school closure, cancel public events, international and local travel ban, etc), financial response (income support, debt relief for households, etc), health systems (emergency investment in healthcare, contact tracing, etc), and miscellaneous responses. These indicators are used to compare government responses and measure their effect on the rate of infections. A global map of the world ranking countries with the Stringency Index is presented. Data is collected from internet sources, news articles, and government press releases. Data is available in multiple formats and interfaces on the team website and GitHub repository ^43^. The project is titled Oxford COVID-19 Government Response Tracker (OxCGRT) and has a working paper and corresponding open source repository.

A similar data set of 117 countries has been curated by a group of volunteers ^44^. The objective of the data set curation is to study the effectiveness of NPI on national scales without consideration of economic stimulus. The data set includes information on lock-down measures, travel bans, and testing counts. The authors acknowledge the sampling bias in the data set as some countries are difficult to document and the government reports may differ from actual implementation and consequences. The data set can be utilized to study the correlation between national responses and infection transmission rates.

ACAPS, an independent information provider, details a dashboard ^45^ and CSV files ^46^ for COVID-19 government measures which is updated weekly. The collected information falls into the categories of social distancing, movement restrictions, public health measures, social and economic measures, and lock-downs. Each information regarding government measures is elaborated with country, administration level, region, implementation date, and source among other details.

## 5 Speech data sets

The speech (audio) data sets help in COVID-19 diagnosis and detection through three basic methods. Firstly, cough sounds can help in detecting a COVID-19 positive case after the application of ML techniques [7, 36]. Secondly, breathing rate can be detected from speech resulting in the screening of a person for COVID-19[95]. Thirdly, stress detection techniques from speech can be used to detect persons with indications of mental health problems and the severity of COVID-19 symptoms. All these techniques require extensive efforts for data set collection. These speech-based COVID-19 diagnosis techniques can be enabled by smartphone applications or remote medical care through telemedicine.

### 5.1 Cough based COVID-19 diagnosis

Imran et al. [7] exploited the fact that cough is one of the major symptoms of COVID-19. What makes this exploitation process complex, is the truth that cough is a symptom of over thirty non-COVID-19 related medical conditions. To address this problem, the authors investigate the distinctness of pathomorphological alterations in the respiratory system induced by COVID-19 infection when compared to other respiratory infections. Transfer learning is exploited to overcome the COVID-19 cough training data shortage. To reduce the misdiagnosis risk stemming from the complex dimensionality of the problem, a multi-pronged mediator centered risk-averse AI architecture is leveraged. The AI architecture consists of three independent classifiers, i.e., Deep Learning-based multi-class classifier, Classical ML-based multi-class classifier, and Deep Learning-based binary class classifier. If the output of any classifier mismatches other, inconclusive result is returned. Results show that proposed AI4COVID-19 can distinguish among COVID-19 coughs and several types of non-COVID19 coughs. The accuracy of more than 90% is promising enough to encourage a large-scale collection of labeled cough data to gauge the generalization capability of AI4COVID-19.

Researchers [37] developed a cross-platform application for crowd-sourced collection of voice sounds (cough and breath) to distinguish healthy and unhealthy persons. The voice sounds are used to distinguish between COVID-19, asthma, and healthy persons. Three binary classification tasks are constructed i.e., **(a)** distinguish COVID-19 positive users from healthy users **(b)** distinguish COVID-19 positive users who have a cough from healthy users who have a cough, and **(c)** distinguish COVID-19 positive users with a cough from users with asthma who declared a cough. More than 7000 unique users (approximately 10K samples) participated in the crowdsourced data collection out of which more than 200 reported being COVID-19 positive. Standard audio augmentation methods were used to increase the sample size of the data set. Three classifiers, namely, Logistic Regression, Gradient Boosting Trees, and SVM were utilized for the classification task. The study utilizes the aggregate measure of the area under the curve (AUC) for performance comparison. AUC of greater than 70% is reported in all three binary classification tasks. The authors also utilize breathing samples for classification and find AUC to be approximately 60%. However, when the cough and breathing inputs are combined for classification, the AUC improves to approximately 80% for each task due to a higher number of features.

Sharma et al. [96] aim to supplement the laboratory-based COVID-19 diagnosis methods with cough based diagnosis. The project, named is Coswara, utilizes cough, breath, and speech sounds to quantify biomarkers in acoustics. Nine different vocal sounds are collected for each patient including breath (shallow and deep), cough (shallow and heavy), and vowel phonations. The nine vocals capture different physical states of the respiratory system. Multi-dimensional spectral and temporal features are extracted from audio files. The classification and data curation tasks are under process.

The work is supplemented by a web application for data collection ^47^ and open source voice data set of approximately 1000 samples in wav format (44.1KHz) ^48^.

In summary, detecting/screening COVID-19 from cough samples using ML techniques has indicated promising results [7, 36]. The accuracy of the studies is hindered due to the small data set of COVID-19 cough samples. Several researchers are gathering cough based data and have made appeals for contribution from the public. Researchers from the University of Cambridge ^49^, Carnegie Mellon University ^50^, and EPFL ^51^ have made calls for the community participation in the collection of the cough data. An independent AI team has made the call for data collection ^52^ and also made an open source repository for the cough data ^53^. However, the data set consists of only 16 samples (7 COVID-19 PCR test positives, 9 negative). Further efforts are required towards data collection to enable the application of ML techniques for higher accuracy diagnosis. Figure 4 depicts a work-flow of speech-based COVID-19 diagnosis [96, 7].

**Figure 4:**
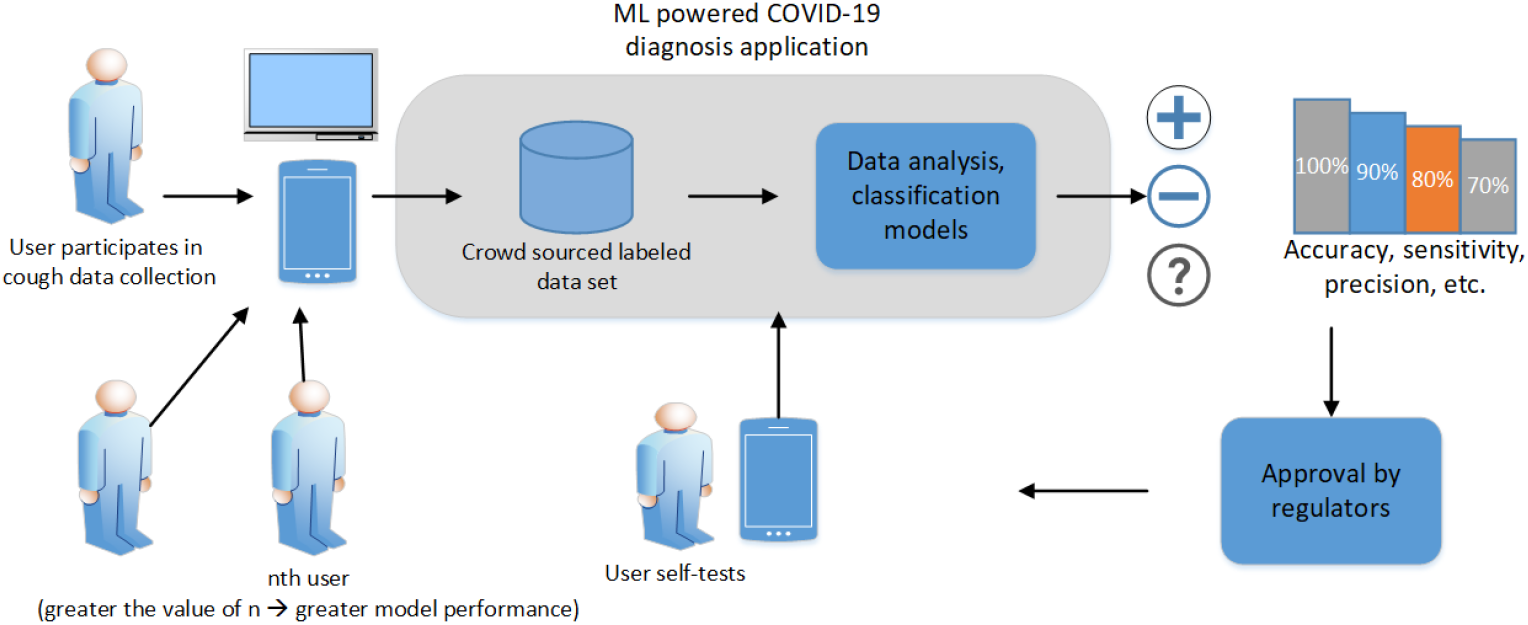
A work-flow of speech based COVID-19 diagnosis.

### 5.2 Breath based COVID-19 diagnosis

Breathlessness or shortness of breath is a symptom in nearly 50% of the COVID-19 patients which can also indicate other serious diseases such as pneumonia [95]. Automated detection of breathlessness from the speech is required in remote medical care and COVID-19 screening applications. Patient speech can be recorded for breath patterns with a simple microphone attached to smart devices. Abnormality related to COVID-19 can be detected from the breath patterns.

Faezipour et al. [97] proposed an idea smartphone application for self-testing of COVID-19 using breathing sounds. The authors imply that breathing difficulties due to COVID-19 can reveal acoustic patterns and features necessary for COVID-19 pre-diagnosis. The breathing sounds can be input to the smartphone through the microphone. Signal processing, ML, and deep learning techniques can be applied to the breathing sound to extract features and classify the input into COVID-19 positive and negative cases. Such a smartphone-based application can be used as a self-test while eliminating the risks and costs associated with visiting medical facilities. The proposed framework can be augmented with data obtained from a spirometer (lung volume) and blood oxygenation measured from a pulse oximeter. The data should be initially labeled as COVID-19 positive and negative by medical experts based on clinical findings to train the proposed model. Afterward, ML techniques can extract features and classify new inputs based on model training.

Authors [98] detailed a portable smartphone powered spirometer with automated disease classification using CNN. Spirometer is a device that measures the volume of expired and inspired air. The proposed system consists of three basic modules. First, fleisch type airflow tube captures the breath with a differential pressure-based approach. A blue-tooth enabled micro-controller is built for data processing. Lastly, an Android application with a pre-trained

CNN model for classification is developed. Stacked AutoEncoders, Long Short Term Memory Network, and CNN are evaluated as classifiers for lung diseases such as obstructive lung diseases and restrictive lung diseases. The 1-D CNN classifier exhibits higher accuracy than other ML classifiers. The proposed model can be extended to include COVID-19 classification and the classifiers can be re-evaluated for accuracy. In summary, tools are available for the breath-based COVID-19 diagnosis. However, existing applications are required to be updated accompanied by voice collections from COVID-19 patients.

### 5.3 Speech based COVID-19 severity estimation

Han et al. [99] provide an initial data driven study towards speech analysis for COVID-19 and detect physical and mental states along with symptom severity. Voice data from 52 patients hospitalized in China was gathered with five sample sentences. Moreover, each patient was asked to rate his sleep quality, fatigue, and anxiety on low, average and high level. Demographic information for each patient was also collected. The data was pre-processed in four steps namely, data cleansing, hand annotating of voice activities, speaker diarisation, and speech transcription. openSMILE toolkit was used to extract two feature sets namely, Computational Paralinguistics Challenge (COMPARE) and the extended Geneva Minimalistic Acoustic Parameter (eGeMAPS). Four classification tasks are performed on data. Firstly, the patient severity is estimated with the help of number of hospitalization days. The rest of the three classification tasks predict the severity of self-reported sleep quality, fatigue, and anxiety levels of COVID-19 patients with SVM classifier and linear kernel. Performance in terms of unweighted average recall showed promising results for sleep quality and anxiety prediction.

## 6 Comparison

In this section we provide a tabular and descriptive comparison of the surveyed open source data sets. Table 1 presents the comparison of medical image data sets in terms of application, data type, and ML method in tabular form.

**Table 1:**
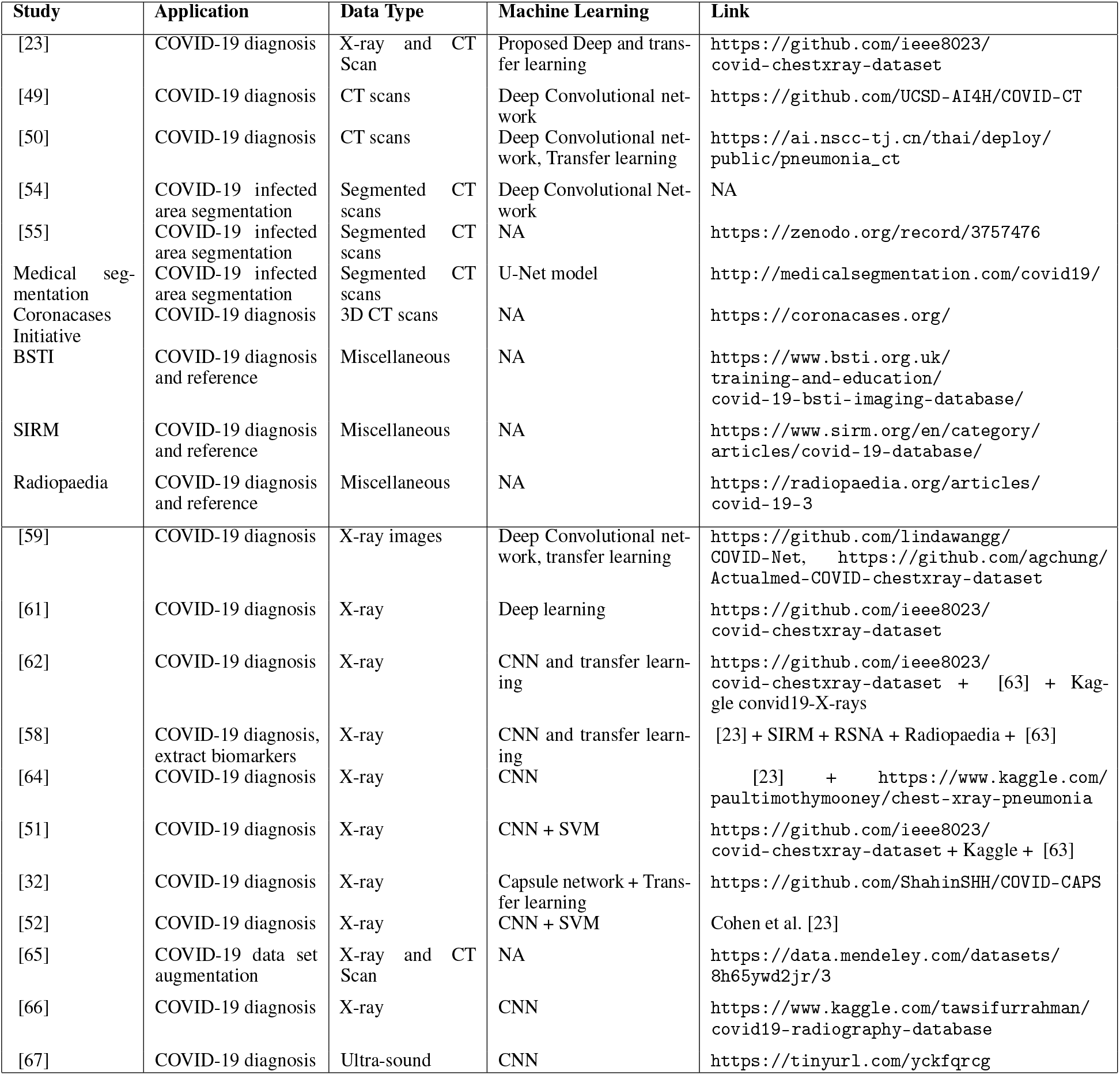
Comparison of COVID-19 medical image data sets

First, we compare all of the listed works on their openness. Some of the works do not have data and code publicly available and it is difficult to validate their work [100]. Others have code or data publicly available [101]. Such studies are more relevant in the current pandemic for global actions concerning verifiable scientific research against COVID-19. On the other hand, some studies merge multiple data sets and mention the source of data but do not host it as a separate repository [102]. The highly relevant studies have made public both data and code [49, 42].

Higher number of reported works have utilized X-ray images than CT scans. Very few studies have utilized ultrasounds and MRT images [67]. Segmentation techniques to identify infected areas have been mostly applied to CT scans [54]. Similarly, augmentation techniques to increase the size of the data set have been applied mostly to X-ray image based data sets [52, 65]. All of the works provided 2D CT scans except for one resource from the Coronacases Initiative ^54^. Most of the COVID-19 diagnosis works employed CNNs for classification. Some of the works utilized transfer learning to further increase the accuracy of classification by learning from similar tasks [50, 58]. Moreover, few works augmented CNNs with SVM for feature extraction and classification tasks [52, 51]. Higher accuracies were reported from works augmenting transfer learning and SVM with CNNs. CNNs and deep learning techniques are reported to overfit models due to the limited size of COVID-19 data sets [49]. Therefore, authors also researched alternative approaches in the form of Capsule network [32] and SVM [51] for better classification on limited data sets of COVID-19 cases. Augmentation techniques have also been employed to increase the size of data set. However, further analysis is required on the performance evaluation of COVID-19 diagnosis on augmented data sets [65].

Most of the COVID-19 diagnosis works distinguished between two outcomes of COVID-19 positive or negative cases [64]. However, some of the works utilized three outcomes, i.e., COVID-19 positive, viral pneumonia, and normal cases for applicability in real-world scenarios. Researchers [58] expanded the classification to six common types of pneumonia. Such methodologies require the extraction and compilation of data sets with other categories of pneumonia radiographs. The ML-based COVID-19 diagnosis is difficult to fully automate as a human in the loop (HITL) is required to label radiographic data [54]. Segmentation techniques have been utilized to embed bio-markers in data set [55]. However, the segmentation techniques also require HITL for verification.

ResNet, MobileNet, and VGG have been commonly employed as pre-trained CNNs for classification [61, 62]. AI/ML explainability methods have been seldom used to delineate the performance of CNNs [60]. Most of the works report accuracies greater than 90% for COVID-19 diagnosis [52, 51]. The data sets of Cohen et al. [23] is considered pioneering effort and is mostly utilized for the COVID-19 cases and Kermany et al. [63] is employed for common pneumonia cases.

Table 2 presents the comparison of textual data sets (COVID-19 case reports) in terms of application, data type, and statistical method in tabular form.

**Table 2:** Comparison of COVID-19 case report data sets

| Study | Application | Data Type | Statistical method | Link |
| --- | --- | --- | --- | --- |
| [43] | Reporting global cases | COVID-19 cases | NA | https://github.com/CSSEGISandData/COVID-19 |
| [70] | COVID-19 visual analysis | COVID-19 statistics | Exploratory data analysis | WHO + John Hopkins + Chinese Center for Disease Control and Prevention |
| [71] | COVID-19 city wise case analysis in China | COVID-19 statistics | NA | https://github.com/cheongsa/Coronavirus-COVID-19-statistics-in-China |
| [19, 72] | Reporting China cases | Location and epidemiological data | NA | https://github.com/beoutbreakprepared/nCoV2019/tree/master/latest_data |
| [42] | US county level data | 348 socioeconomic parameters | proposed ML for epidemiological analysis | https://github.com/JieYingWu/COVID-19_US_County-level_Summaries |
| [4] | Estimating new cases | COVID-19 cases | stochastic transmission dynamic | https://github.com/adamkucharski/2020-ncov/ |
| [73] | COVID-19 spread | COVID-19 statistics | ARIMA | https://github.com/CSSEGISandData/COVID-19 |
| [74] | Correcting under-reported cases | Reported case and world demographics | Statistical | https://tinyurl.com/y7hbp196 |
| [44] | Mobility-transmission analysis | Mobility and epidemiological data | Statistical | https://github.com/Emergent-Epidemics/covid19_npi_china |
| [75] | Cases exported from China | epidemiological data set | Statistical | http://www.mdpi.com/2077-0383/9/2/601/s1 |
| [76] | Effect of NPI on COVID-19 in China | Location and epidemiological data | NA | https://github.com/wpgp/BEARmod |
| [45] | Effect of NPI on COVID-19 in Europe | Location and epidemiological data | semi-mechanistic Bayesian hierarchical model | https://github.com/ImperialCollegeLondon/covid19model/releases/tag/v1.0 |
| [77] | International travel control analysis | COVID-19 statistics, flight data | Statistical | https://github.com/WellsRC/Coronavirus-2019 |
| [78] | COVID-19 Transmission control analysis | COVID-19 statistics | regression analysis | https://github.com/huaiyutian/COVID-19_TCM-50d_China |
| [79] | Community transmission | COVID-19 cases | Expectation-maximization | https://github.com/carolinecolijn/ClustersCOVID19 |
| [80] | Community transmission | COVID-19 cases (dates) | maximum likelihood fitting and the Akaike information criterion | https://github.com/MeyersLabUTexas/COVID-19 |
| [81] | Community transmission | COVID-19 cases (dates) | Bayesian approach | https://github.com/aakhmetz/nCoVSerialInterval2020 |

The textual data sets are applied for multiple purposes, such as, **(a)** reporting and visualizing COVID-19 cases in time-series formats [43, 4], **(b)** estimating community transmission [81], **(c)** correlating the effect of mobility on virus transmissions [44], **(d)** estimating effect of NPI on COVID-19 cases [76, 45], **(e)** forecasting reproduction rate and serial intervals, **(f)** learning emotional and socio-economic issue from social media [82, 84], and **(g)** analyzing scholarly publications for semantics [91]. Most of the articles apply statistical techniques (stochastic, Bayesian, and regression) for estimation and correlation of data [45, 73]. There is great scope for the application of AI/ML technique as proposed in some studies [5, 42]. However, only statistical techniques have been applied to textual data sets in most of the listed works. Most of the studies that estimate COVID-19 transmissions utilize COVID-19 case data collected from various governmental, journalistic, and academic sources [42]. The case reports are available in visual dashboards and CSV formats.

Table 3 presents the comparison of textual data sets (social media and scholarly articles) in terms of application, data type, and statistical method in tabular form. The studies that analyze human emotions have mostly utilized Twitter API to collect data [85, 5]. Studies estimating effect of NPI on COVID-19 bring to service location, mobility, epidemiological, and demographic data [45]. The collection of scholarly articles have proposed potential of data science and NLP techniques [6] while a demonstration of the same is available at COVIDScholar. However, the details of the semantic analysis algorithms applied by the COVIDScholar are not available. Github is the first choice of researchers to share open access data while Kaggle is seldom put to use [82, 64].

**Table 3:**
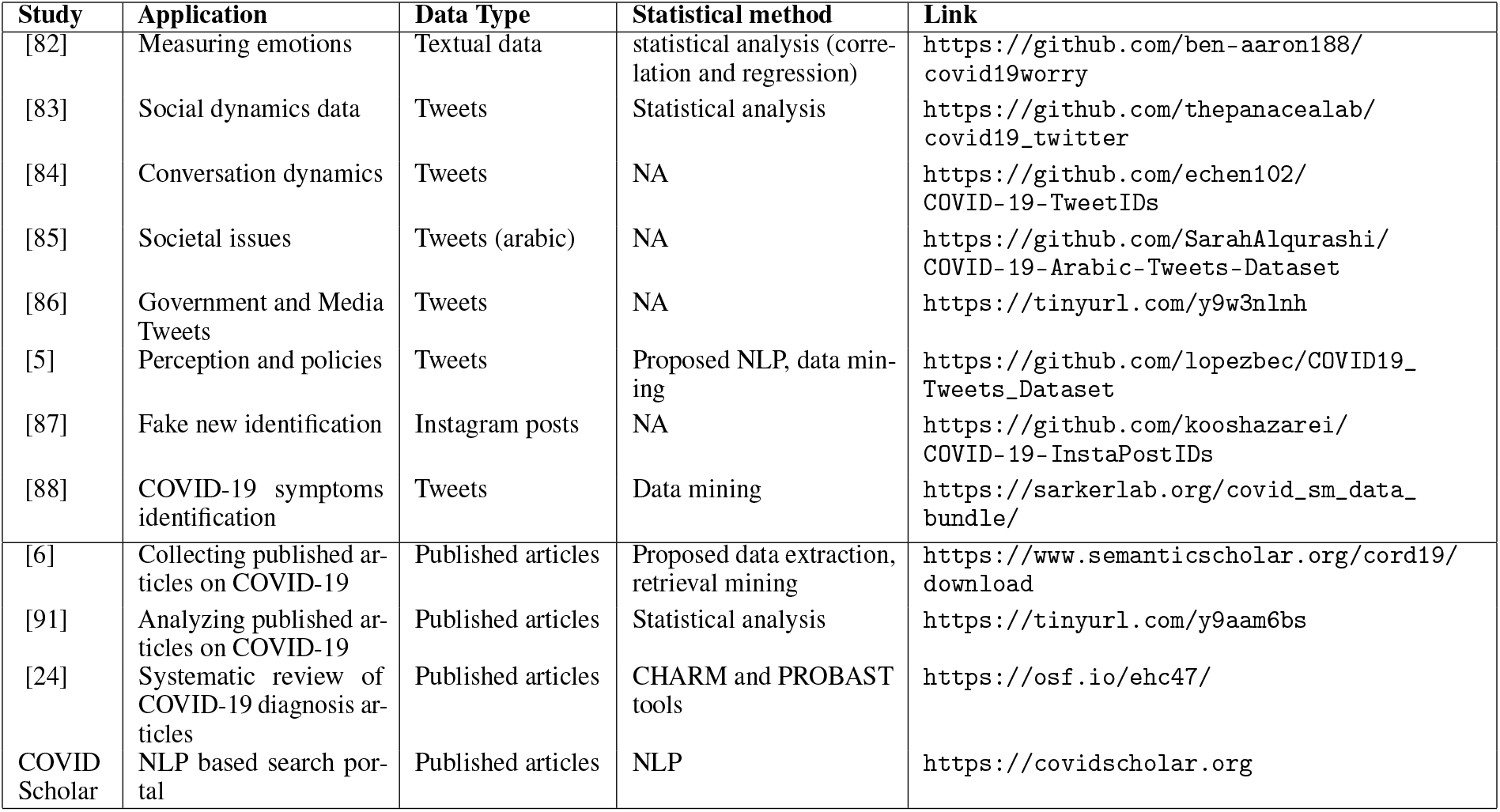
Comparison of COVID-19 social media and scholarly article data sets

Table 4 provides a comparison of mobility and NPI data sets based on parameters including data set application, source, format, and coverage. Mobility data sets provided by Google, Apple, and Baidu aid the analysis of COVID-19 case transmissions [78, 44]. The mobility data is collected by location services provided on smart devices. The mobility data is usually anonymised with random identifiers to keep user privacy intact. However, the privacy measures of Baidu are not known. Several efforts have been made to record NPI at the country level from information released by media and government press [94]. The NPI data in conjunction with infection rates can shed light on the effectiveness of the government policies. The mobility and NPI data sets are available in dashboard info-graphics and in CSV format.

**Table 4:**
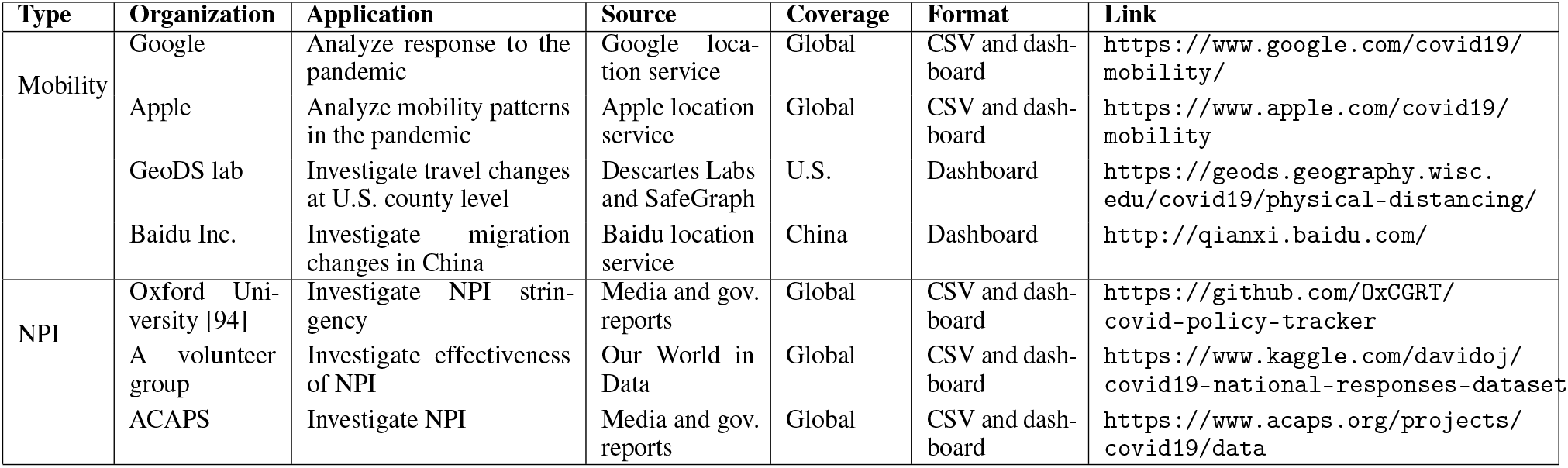
Comparison of COVID-19 Mobility and NPI data sets

Table 5 lists the speech data based research works for COVID-19 in terms of application, data type, ML methods, and data set size. The applications of speech data for COVID-19 diagnosis are very encouraging as identified in most of the listed research works in section 4.4. Most of the listed works focus on cough based COVID-19 diagnosis from speech data. ML classifiers such as SVM and CNN are commonly utilized for classification. Although there are multiple mobile applications for collection of voice data, open source data sets are few and very small in size. Further open source data collection is required for **(a)** application of deep learning methods, **(b)** application of methods for COVID-19 severity prediction, and **(c)** prediction of patient behavioral features (mental health, anxiety, stress, etc) from speech data [103, 36].

**Table 5:**
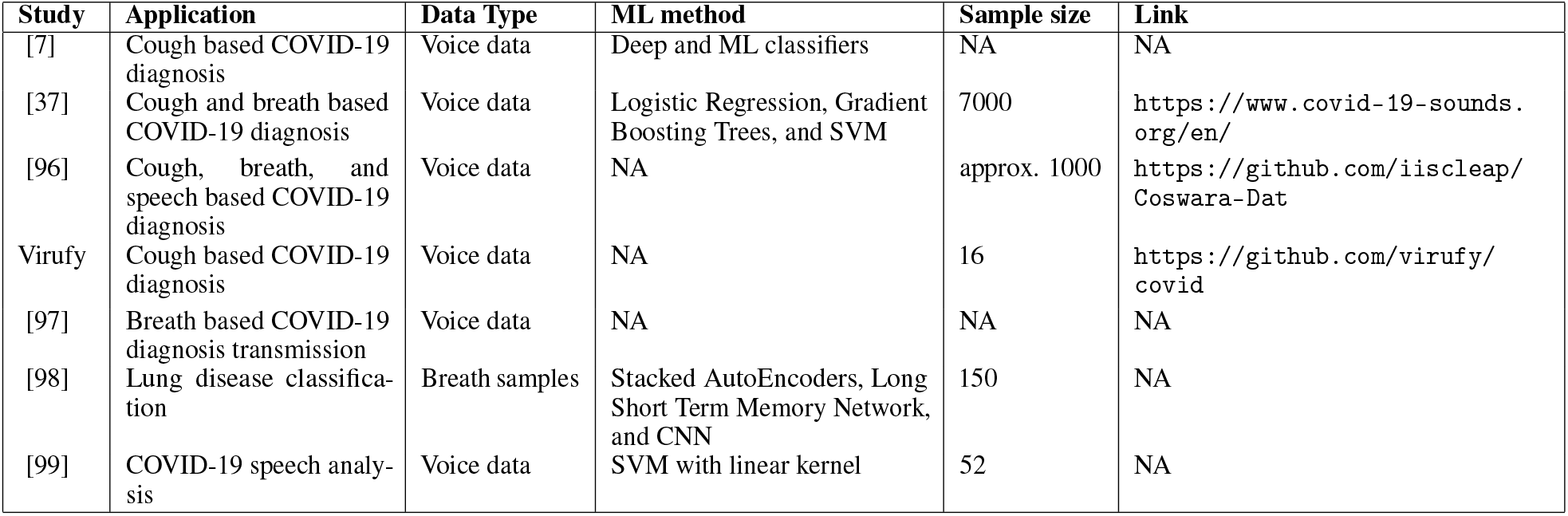
Comparison of COVID-19 Speech data sets

| Study | Application | Data Type | ML method | Sample size | Link |
| --- | --- | --- | --- | --- | --- |
| [7] | Cough based COVID-19 diagnosis | Voice data | Deep and ML classifiers | NA | NA |
| [37] | Cough and breath based COVID-19 diagnosis | Voice data | Logistic Regression, Gradient Boosting Trees, and SVM | 7000 | https://www.covid-19-sounds.org/en/ |
| [96] | Cough, breath, and speech based COVID-19 diagnosis | Voice data | NA | approx. 1000 | https://github.com/iiscleap/Coswara-Dat |
| Virufy | Cough based COVID-19 diagnosis | Voice data | NA | 16 | https://github.com/virufy/covid |
| [97] | Breath based COVID-19 diagnosis transmission | Voice data | NA | NA | NA |
| [98] | Lung disease classification | Breath samples | Stacked AutoEncoders, Long Short Term Memory Network, and CNN | 150 | NA |
| [99] | COVID-19 speech analysis | Voice data | SVM with linear kernel | 52 | NA |

Most of the research on COVID-19 is currently not peer-reviewed and in the form of pre-prints. The COVID-19 pandemic is a matter of global concern and necessitates that any scientific work published should go through a rigorous review process. At the same time, the efficient diffusion of scientific research is also demanded. Therefore, this review had to include pre-prints that have not been peer-reviewed to compile a comprehensive list of articles. The pre-prints contribute approximately 50% of the cited research in this review. The credibility of reviewed work is supported by the open source data sets and code accompanying the pre-prints. The research works can be compared on the re-usability metric of the data sets such as Meloda 5 [104, 105].

## 7 Discussions and Future Challenges

There are multiple challenges to AL/ML-based COVID-19 research and data. The foremost on the list is the availability and openness of data [29, 30, 31]. As more open source data is made available, AI/ML-based research collaborations across the globe, system verification, and real-world operations will be possible. We detail the future challenges in the following subsection.

### 7.1 Open source data

The AI/ML techniques are often open source and implemented as libraries and packages in programming language development platforms. Some of the examples are Scikit-learn module in Python [10] and Weka library in R [106]. As a result, the focus shifts towards openness and availability of data. The novel COVID-19 pandemic necessitates creation, management, hosting, and benchmarking of new data sets. Existing research works lean more towards opaque research methodology rather than open source methods. Each of these practices has its pros and cons. The closed source research can lead towards patenting of research innovations and ideas. It can also lead to collaboration in closed research groups across the globe. However, withholding critical data in the context of COVID-19 may be considered maleficence [30]. Moreover, open source research methods offer greater benefits that are more far-reaching while accelerating AI/ML innovations for the COVID-19 pandemic. The abrupt spread of the COVID-19 pandemic has also highlighted the open source data as the current key barrier towards AI/ML-based combat against COVID-19 [21]. We list points below on future challenges of open source data sets.

- Most of the data and code on COVID-19 analysis is closed source. Whatever data is available, it is limited for applications of deep learning methods. Efforts to curate and augment existing data sets with samples from hospitals and clinics (medical data sets) and self-testing (cough and breath data) applications are specifically required [7, 28].
- Data should be created, managed, hosted, processed, and bench-marked to accelerate COVID-19 related AI/ML research. As more data is integrated, the (deep) learning techniques become more accurate and move towards large-scale operations. Labeling of large data sets is another indispensable task [28, 107].
- The scarcity of the data is attributed to **(a)** closed source research methods, **(b)** distributed nature of data (medical images may be available at a hospital but not aggregated in a unified database), and **(c)** privacy concerns limiting data sharing. Therefore, a key challenge is the federation of the data sets to combat AI. Standards and protocols along with international collaborations are necessary for the federation of data sets. The privacy concerns can be addressed by adopting standard procedures for anonymity of the data [20, 13].
- Interpretability and explainability of AI/ML techniques is another key challenge. ML techniques act as a blackbox. Specifically, in deep learning, doctors and radiologists must know which features distinguish a COVID-19 case from non-COVID-19. Moreover, the probability of error needs to be estimated and communicated with the practitioners and patients [107, 30].
- Most of the medical data comes from China and European countries which may lead to selection bias when applied in other countries. As a result, the practice of diagnosing a patient with COVID-19 using AI/ML is very rare. Moreover, it is yet to be investigated if AI/ML can detect COVID-19 before its symptoms appear in other laboratory methods to justify its practice [30].

### 7.2 Challenges to medical data

Most of the researchers studying image-based diagnoses of COVID-19 have emphasized that further accuracy is required for the application of their methods in clinical practices. Moreover, researchers have also emphasized that the primary source of COVID-19 diagnosis remains the RT-PCR test and medical imaging services aim to aid the current shortage of test kits as a secondary diagnosis method [50, 108, 30]. Contact-less work-flows need to be developed for AI-assisted COVID-19 screening and detection to keep medical staff safe from the infected patients [13, 109]. A patient with RT-PCR test positive can have a normal chest CT scan at the time of admission, and changes may occur only after more than two days of onset of symptoms [108]. Therefore, further analysis is required to establish a correlation between radiographic and PCR tests [110].

Data sets are available for most of the research directions in biomedical imaging. However, these data sets are limited in size for the application of deep learning techniques. Researchers have emphasized that larger data sets are required for deep learning algorithms to provide better insights and accuracy in diagnosis [52]. Therefore, the collaboration of medical organizations across the globe is necessary for expanding existing data sets. Moreover, the accuracy of augmentation techniques in increasing the data set size needs to be evaluated. The CT scan and X-ray based data sets and research are conventional. MRI provides high-resolution images and soft-tissue contrast at a higher cost. MRI based COVID-19 diagnosis and data set are demanded to compare their accuracy with CT-scans and X-ray based methods. Moreover, the operational performance and effectiveness of the proposed AI/ML-based diagnosis in clinical work-flows under regulatory and quality control measures and unbiased data needs investigation [28, 111].

The research on speech-based diagnosis of COVID-19 on symptoms of cough and breath rate is in the early stage of development. Researchers have made calls for the collection of voice data, However, whatever data is utilized in the existing studies, only one open source data sets is available yet for speech based COVID-19 diagnosis. Moreover, the existing data set sizes need enhancement for higher accuracy of classification tasks.

### 7.3 Challenges to textual data

Three reported studies collected scholarly articles related to COVID-19 [91, 6, 24]. However, the application of NLP is proposed in these works. The inference of scientific facts from published scholarly articles remains a challenge yet to be addressed in reference to COVID-19. The only resource available in this direction is the COVIDScholar developed by Berkeley Lab for semantic analysis of COVID-19 research. However, the details of their algorithms are not available. Similarly, data sets have been curated from social media platforms [85, 5]. The human emotions and psychology in the pandemic, sentiments regarding lock-downs, and other NPI are yet to be investigated thoroughly. Another research direction is related to social distancing in the pandemic. Given the preferred social distance across multiple countries and the open source data on mobility, what are the effects on COVID-19 transmission? [112]. Moreover, data set curation is also needed to provide an update on practiced social distances during the COVID-19 lock-down initiatives. The timeliness of the research is another important issue for textual data. Social media data analysis and corresponding actions become outdated very quickly as it is collected, pre-processed, and annotated at large-scale [28].

### 7.4 Privacy issues

As user’s data regarding mobility, location, medical diagnosis, and social media is utilized in ML and statistical studies, privacy remains a focal issue. Privacy concerns are further escalated due to open source nature of data. Privacy concerns can dominate public health concerns leading to limited sharing of data for scientific purposes. Moreover, there is a fear that mission creep will occur when this pandemic is over and the governments will keep on tracking and surveying populations for other purposes. Users have concerns about large-scale governmental surveillance in case such data from applications is shared with a third party [30, 113]. Google, Apple, Baidu, and SafeGraph have been identified as sources for mobility data in this review. Similarly, hospitals and medical organizations have contributed to the collection of medical data. Efforts have been made on the anonymity of the data. However, the data sets have not been rigorously tested for security and privacy vulnerabilities.

The automated contact tracing application initiated by several governments in the wake of COVID-19 transmissions also demands consideration of user privacy issues [114]. Automated contact tracing applications monitor the user-user interactions with the help of Bluetooth communications. The population at risk can be identified if one user is diagnosed as COVID-19 positive from his user-user interactions automatically saved by the contact tracing application [115]. The contact tracing applications can be utilized for large-scale surveillance as user data is updated in a central repository frequently. It is yet to be debated the compliance of contact tracing applications with country-level health and privacy laws. Similarly, patient privacy concerns need to be addressed on the country level based on health and privacy laws and social norms [116].

Public hatred and discrimination have also been reported against COVID-19 patients and health workers [33]. The situation demands complete anonymity of medical and mobility data to avoid any discrimination generating from data shared for academic purposes. Blockchain and federated learning are two perspective solutions for additional privacy measures. Private Blockchain can provide privacy and accountability for data access as it is able to trace the data operations with trust and decentralization features [26, 117]. Several blockchain-based privacy-preserving solutions have been proposed in the context of COVID-19 pandemic [118, 113]. Federated learning-based ML techniques do not need to share and store data at a centralized location such as a cloud data center. ML models are distributed over participating nodes while sharing only model parameters and outputs with the central node. As a result, privacy is preserved in a federated framework of machine learning [36, 119]. Moreover, public health may be prioritized over individual privacy issues in the context of COVID-19 pandemic [28].

### 7.5 Misuse of Digital Technologies

Although digital technologies have significantly aided the combat against COVID-19, they have also provided the d for vulnerabilities that can be exploited in terms of social behaviors [120]. Fake news/misinformation sharing cial media platforms [15], racist hatred [16], propaganda (against 5G technologies and governments) [17], and e financial scams [18] are few forms of digital platform exploitation in COVID-19 pandemic. Fake news and rumors have been spread about lock-downs policies, over-crowded places, and death cases on social media platforms.-news identification is already a popularly debated topic among the social and data science community [121]. ing NLP techniques for fake news identification need to be applied on COVID-19 social media data sets for ation of proposed works in the current pandemic [122, 16]. The focus needs to be on the timely identification ke news as with increasing time, all actions may become irrelevant. The social media platforms also need to be zed for human perceptions and sentiments regarding specific ethnicity (for example, sinophobia) and lock-down ies [123]. Alarming rise in hate speech and misinformation has been reported during the pandemic on the Internet. ropaganda that 5G networks are responsible for COVID-19 spread has also received widespread attention on social media. With the society heavily relying on online shopping and banking transactions in the pandemic, an increased er of online scamming and hacking activities have been reported [18]. Therefore, it is necessary to address and ate the misuse of technologies while we heavily rely on them in the existing pandemic for information, retail, tainment, and banking.

Other future challenges related to the theme of our article are, but not limited to, **(a)** forecasting COVID-19 cases atalities on city and county levels, **(b)** predicting transmission factors, incubation period, and serial interval on munity level, **(c)** using NLP to analyze public sentiments regarding COVID-19 policies from social media, **(d)** ing NLP on scholarly articles to automatically infer scientific findings regarding COVID-19, **(e)** identifying key (obesity, air pollution, etc) and social risk factors for COVID-19 infections, **(f)** identifying demographics at more f infection from existing cases and trends, and **(g)** ethical and social consideration of analyzing patients data. from these, multiple challenges and competitions are being hosted on Kaggle to address issues pertinent to open e COVID-19 data sets ^55, 56, 57^ and elsewhere on the Internet ^58^.

## 8 Conclusion

We provided a comprehensive survey of COVID-19 open source data sets in this article. The survey was organized e basis of data type and data set application. Medical images, textual data, and speech data formed the main ypes. The applications of open source data set included COVID-19 diagnosis, infection estimation, mobility and graphic correlations, NPI analysis, and sentiment analysis. We found that although scientific research works on COVID-19 are growing exponentially, there is room for open source data curation and extraction in multiple directions as expanding of existing CT scan data sets for application of deep learning techniques and compilation of data set ugh samples. We compared the listed works on their openness, application, and ML/statistical methods. Moreover, we provided a discussion of future research directions and challenges concerning COVID-19 open source data sets. We that the main challenge towards data-driven AI is the opaqueness of data and research methods. Further work is red on **(a)** the curation of data set for cough based COVID-19 diagnosis, **(b)** expanding CT scan and X-ray data sets gher accuracy of deep learning techniques, **(c)** establishment of privacy-preserving mechanisms for patient data, mobility, and contact tracing, **(d)** contact-less diagnosis based on biomedical images to protect front-line health workers from infection, **(e)** sentiment analysis and fake new identification from social media for policy making, and **(f)** ntic analysis for automated knowledge-based discovery from scholarly articles to list a few. We advocate that the works listed in this survey based on open source data and code are the way forward towards extendable, transparent, and verifiable scientific research on COVID-19.

## Data Availability

We provide direct downloadable links for most of the datasets discussed in the manuscript

## Aknowledgments

This work was supported by the Research and Development Office, Ministry of Education, Saudi Arabia.

## Footnotes

2 https://www.bbc.com/news/business-52483082

3 https://ai.nscc-tj.cn/thai/deploy/public/pneumonia_ct

4 https://zenodo.org/record/3757476

5 https://gitee.com/junma11/COVID-19-CT-Seg-Benchmark

6 http://medicalsegmentation.com/covid19/

7 https://www.sirm.org/en/category/articles/covid-19-database/

8 https://coronacases.org

9 https://www.bsti.org.uk/training-and-education/covid-19-bsti-imaging-database/

10 https://www.sirm.org/en/category/articles/covid-19-database/

11 https://radiopaedia.org/articles/covid-19-3

12 https://www.kaggle.com/andrewmvd/convid19-X-rays

13 https://www.kaggle.com/paultimothymooney/chest-xray-pneumonia

14 Kaggle. https://www.Kagglee.com/andrewmvd/convid19-X-rays

15 https://www.kaggle.com/paultimothymooney/chest-xray-pneumonia

16 https://www.kaggle.com/tawsifurrahman/covid19-radiography-database

17 https://tinyurl.com/yckfqrcg

18 https://pocovidscreen.org/

19 https://www.kaggle.com/covid-19-contributions

20 https://github.com/WeileiZeng/Open-Source-COVID-19

21 https://www.arcgis.com/apps/opsdashboard/index.html

22 https://github.com/CSSEGISandData/COVID-19

23 https://datahub.io/core/covid-19

24 https://ncov.dxy.cn/ncovh5/view/pneumonia

25 https://coronavirus.jhu.edu/map.html

26 https://dataverse.harvard.edu/dataverse/2019ncov

27 https://www.kaggle.com/lachmann12/world-population-demographics-by-age-2019

28 https://www.kaggle.com/lachmann12/correcting-under-reported-covid-19-case-numbers

29 https://github.com/Emergent-Epidemics/covid19_npi_china

30 https://www.ecdc.europa.eu/en/covid-19-pandemic

31 http://qianxi.baidu.com/

32 https://github.com/BayesForDays/coronada

33 https://www.kaggle.com/smid80/coronavirus-covid19-tweets

34 https://covidscholar.org

35 https://www.google.com/covid19/mobility/

36 https://www.kaggle.com/chaibapat/google-mobility

37 https://www.apple.com/covid19/mobility

38 https://geods.geography.wisc.edu/covid19/physical-distancing/

39 https://www.safegraph.com/

40 https://www.descarteslabs.com/

41 http://qianxi.baidu.com/

42 https://flirt.eha.io/

43 https://github.com/OxCGRT/covid-policy-tracker

44 https://www.kaggle.com/davidoj/covid19-national-responses-dataset

45 https://www.acaps.org/projects/covid19/data

46 https://tinyurl.com/yc2m29vz

47 https://coswara.iisc.ac.in/

48 https://github.com/iiscleap/Coswara-Data

49 https://www.covid-19-sounds.org/en/

50 https://cvd.lti.cmu.edu/

51 https://coughvid.epfl.ch/

52 http://virufy.org/

53 https://github.com/virufy/covid

54 https://coronacases.org/

